# PCNA thermosensitivity underlies an Ataxia Telangiectasia-like disorder

**DOI:** 10.1101/2022.10.20.22281069

**Authors:** Joseph Magrino, Veridiana Munford, Davi Jardim Martins, Thais K Homma, Brendan Page, Christl Gaubitz, Bruna L Freire, Antonio M Lerario, Juliana Brandstetter Vilar, Antonio Amorin, Emília K E Leão, Fernando Kok, Carlos F M Menck, Alexander A L Jorge, Brian A Kelch

## Abstract

Proliferating Cell Nuclear Antigen (PCNA) is a sliding clamp protein that coordinates DNA replication with various DNA maintenance events that are critical for human health. Recently, a hypomorphic homozygous serine to isoleucine (S228I) substitution in PCNA was described to underlie a DNA repair disorder known as PCNA-Associated DNA Repair Disorder (PARD). PARD symptoms range from UV sensitivity, neurodegeneration, telangiectasia, and premature aging. We, and others, previously showed that the S228I variant changes the protein binding pocket of PCNA to a conformation that impairs interactions with specific partners. Here, we report a second PCNA substitution (C148S) that also causes PARD. Unlike PCNA-S228I, PCNA-C148S has WT-like structure and affinity towards partners. In contrast, both disease-associated variants possess a thermo-stability defect. Furthermore, patient-derived cells homozygous for the *C148S* allele exhibit low levels of chromatin-bound PCNA and display temperature-dependent phenotypes. The stability defect of both PARD variants indicates that PCNA levels are likely an important driver of PARD disease. These results significantly advance our understanding of PARD and will likely stimulate additional work focused on clinical, diagnostic, and therapeutic aspects of this severe disease.

## INTRODUCTION

Proliferating Cell Nuclear Antigen (PCNA) is a sliding clamp protein that acts as a molecular “hub” for chromosomal DNA replication, repair, and maintenance. PCNA consists of three identical subunits that form a closed ring that surrounds DNA (1). Each subunit contains one partner binding region that various factors use to interact with PCNA, allowing the trimeric ring to accommodate up to three partners simultaneously (1,2). Because PCNA is a closed ring, it must be loaded onto DNA by the Replication Factor C (RFC) clamp loader complex (3,4). After loading, PCNA functions as a molecular tether that increases the processivity of various DNA-acting enzymes (Figure 1; (5)). Beyond its role in enzymatic processivity, PCNA functions as a scaffold that congregates various factors that copy, surveil, and repair DNA; thus, PCNA coordinates DNA replication with DNA repair, chromatin remodeling, cell cycle regulation, and apoptosis (5). Proper genome stability requires partners to physically interact with PCNA; mutations that abrogate binding can be embryonically lethal (6,7). Therefore, it is critical to understand how PCNA modulates its interactome and elucidate how deviations from normal functional impact human health.

**Figure 1:**
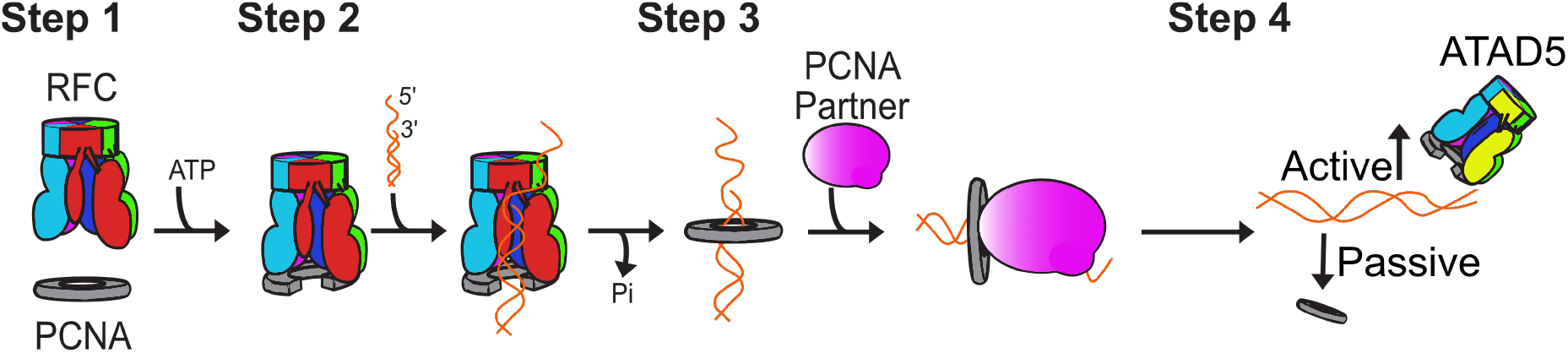
Cartoon representation of PCNA life-cycle. **(Step 1)** The RFC clamp loader binds and loads PCNA onto primer-template DNA in an ATP-dependent manner. **(Step 2-3)** PCNA then remains on DNA, where it interacts with various DNA-interacting enzymes that replicate or maintain the DNA. **(Step 4)** PCNA is removed from DNA either actively by the ATAD5 clamp unloader or by passive dissociation.

Recently, a rare genetic disease, <u>P</u>CNA-<u>A</u>ssociated DNA <u>R</u>epair <u>D</u>isorder (PARD; alternative name: Ataxia-Telangiectasia like disorder 2 (ATLD2)), was reported in four individuals homozygous for the hypomorphic PCNA-S228I variant (8). PARD shares a variety of symptoms (e.g., UV sensitivity, neurological abnormalities, and growth defects) with other DNA repair disorders such as Ataxia Telangiectasia (AT), Cockayne Syndrome (CS), and Xeroderma-Pigmentosum (XP), but presents with a much larger breath of abnormalities (8). Despite its primary role as a replication protein, the S228I variant does not significantly perturb bulk DNA synthesis. In contrast, the S228I substitution increases cells’ sensitivity to UV-induced DNA damage, pointing to a defect in DNA repair and/or the DNA damage response. From a structural standpoint, the S228I substitution dramatically shrinks the partner binding site (9,10), which disrupts interactions between PCNA and a subset of its partners (8–11). Because only certain partner interactions are disrupted by the S228I variant, a view prevailed that PARD is driven by altered specificity for certain binding partners involved in DNA repair (8–11).

In this study, we describe specific clinical and genetic features of three unrelated patients who display apparent PARD symptoms and who are homozygous for a novel *PCNA-C148S* allele. Like PARD patients with PCNA-S228I, these individuals suffer from a combination of photosensitivity, growth delays, and neurodegeneration. Unlike PCNA-S228I cells, cells harboring the C148S variant appear sensitive to dsDNA damaging agents. Furthermore, the C148S alteration does not induce large conformational changes in the PCNA protein, nor does it impair partner binding. However, we find that both variants induce stability defects in the PCNA protein. Furthermore, we observe that patient-derived cells carrying PCNA-C148S exhibit cellular defects consistent with a loss of PCNA stability. Our results challenge the current prevailing idea that altered binding specificity accounts for PARD. We suggest that PCNA longevity and/ or levels of chromatin-bound PCNA underlie this rare disease.

## MATERIALS AND METHODS

### Molecular genetic analysis

This study was approved by the Ethics Committee of Hospital das Clinicas of Sao Paulo University; parental written informed consent was obtained before initiating the genetics studies. DNA samples were extracted from peripheral blood leukocytes using standard procedures. All probands underwent whole-exome sequencing (WES) analyses according to previously published protocols (12,13). Briefly, we constructed an exome library using SureSelect Human All Exon V6 Kit (Agilent Technologies, Santa Clara, USA) following manufacturer’ s instructions. We then sequenced the library on a HiSeq 2500 platform (Illumina, San Diego, USA) using Hiseq SBS V4 cluster generation and sequencing kit (Illumina, San Diego, USA) run on paired-end mode. Reads were aligned to the hg19 assembly of the human genome using the bwa-mem aligner (14). Duplicate reads were flagged with the bammark duplicates tool from biobambam2 which is publicly available through the GNU GENERAL PUBLIC LICENSE Version 3. Variant calling was performed with Freebayes and the resulting Variant Calling Format (VCFs) files were annotated with ANNOVAR (15). The family pedigrees revealed consanguinity among its members; thus, we searched the exome data for the presence of homozygous variants in all patients. Such variants were absent in our in-house sequencing data sets and in public databases (gnomAD, http://gnomad.broadinstitute.org/, and Abraom http://abraom.ib.usp.br/) (16,17). Silent mutations were excluded. The assessment of gene function was performed using the Online Mendelian Inheritance in Man (OMIM) and the PubMed databases. Sanger sequencing was performed to validate the candidate variant identified. The pathogenicity of the C148S variant was then scored using several *in silico* programs: SIFT, PolyPehn2, PROVEAN, CADD, MUtation Assessor, and REVEL (18–23). PCNA sequence alignment were performed using both ClusterOmega and Aminode (24,25).

### Primary fibroblasts and cell culture

Skin primary fibroblasts were obtained from patients 1 and 2. The cells were routinely cultured in DMEM (Gibco ThermoFisher, Waltham, MA, USA or LGC, Cotia, Brazil) supplemented with 10% fetal bovine serum (FBS, Gibco ThermoFisher, USA or Cultilab, Campinas, SP, Brazil) and antibiotics (1% penicillin/streptomycin (Invitrogen, Life Technologies, Carlsbad, CA, USA) in a humidified 5% CO_2_ atmosphere at 37 °C. Healthy donor cells (HDCs) from asymptomatic individuals served as controls (26–28).

### Cell viability with UV irradiation, zeocin treatment, and gamma(γ)-ray irradiation

Cells (5.0 × 10^4^ cells) were seeded in 96 well plates two days prior to treatment and were washed with preheated PBS prior to UV light (260 nm), gamma(γ)-ray, or zeocin treatment. UV dose rates were monitored using a VLX-3W radiometer; 0.1 J/m^2^/s for low-dose exposures and 0.74 J/m^2^/s for high doses. Unirradiated cells were maintained in PBS for the same time as their irradiated counterparts. For γ-ray irradiation, cells were washed with PBS supplemented with 890 μM CaCl_2_ and 500 μM MgCl_2_ and subjected to 1.355 Gy/min using an IBL 637 Cesium-137γ-ray. After UV and γ-ray treatments, cells were incubated with fresh media for 72 hrs. For zeocine treatment (Invitrogen, ThermoFisher), cells were incubated with media supplemented with various concentrations for 72 hrs. Cell viability was performed using the Cellular Proliferation Kit II (XTT, Roche, Basel, Switzerland) according to the manufacturer’ s recommendations. Cell metabolism and viability were assessed in triplicate by the 492 and 650 nm ratio.

### Wound-healing assay

Cells were seeded to 90-100% confluency at 37 ºC 24 hours before the experiment. The monolayers of cells were then scraped with a sterile pipette tip to generate an open wound. Cells were then shifted to either 37 ºC or 42 ºC for 48 hours to allow wound repair. We obtained brightfield images using a EVOS XL Core System microscope (ThermoFisher) and the area of the wound was processed in ImageJ.

### Mutagenesis

Site-directed mutagenesis was performed using untagged PCNA-WT in the pET3c vector to generate PCNA-C148S using 5’ -gcacgtatatc ccgagatctcagccatattg-3’ and 5’ gagatctcggg atatacgtgcaaattcacca-3’ primers (IDT). PCR enzymes for the reaction were purchased from New England BioLabs (NEB, MA USA).

### Protein expression and purification

#### PCNA

All PCNA variants were expressed and purified as previously described (9). Briefly, PCNA variants were expressed from a pET3c vector in *E. coli* BLR-DE3 that were grown in 2XYT supplemented with 100 μg/mL of ampicillin at 37 ºC. Once cells reached an OD600 between 0.6-0.8 they were induced with 1 mM IPTG overnight at 18 ºC. One-liter cultures were centrifuged at 4000 x g and resuspended in Buffer A [25 mM Tris (pH 7.5), 10% (vol/vol) glycerol, and 2 mM DTT], and lysed via cell disruptor (Microfluidics Inc, Westwood, MA.). Lysates were then loaded onto sequential S and 2 x Q 5mL HiTrap columns (GE Healthcare) that were pre-equilibrated with buffer A. S-columns were removed prior to two-column volume washes with Buffer A. PCNA was eluted using a gradient of Buffer B (25 mM Tris (pH 7.5), 10% (v/v) glycerol, 2 mM DTT, and 1 M NaCl). PCNA-containing fractions were further purified via Sephacryl-200 gel filtration column (GE Healthcare) in a Gel Filtration Buffer [20 mM Tris (pH 7.5), 5%(v/v) glycerol, and 2 mM DTT]. PCNA-containing fractions were pooled, concentrated to 20 mg/ml, and flash frozen.

#### N555-hRFC

An N-terminal truncation variant of human RFC was expressed and purified as previously described (11). Briefly, E. coli BLR-DE3 containing p36-p37-p38-p40-pET-Duet and pCDF-1b-RFC-140-N555 were grown in TB media supplemented with 50 µg/mL streptomycin and 100 µg/mL ampicillin at 37 ºC. One-liter cultures were grown to an OD600 ∼ 0.8 then induced with 1 mM IPTG and incubated at 18 ºC overnight. Cells were centrifuged, resuspended in Lysis buffer [20 mM HEPES KOH (pH 7.4), 5% glycerol, 2 mM DTT, 2 mM EDTA, 200 mM NaCl, (w/v), and 0.01% NP-40 (v/v)] supplemented with Mini-Protease Inhibitor Mixture Tablet (Roche), and lysed by cell disruptor. Lysate was loaded onto 5X S columns pre-equilibrated with Buffer A [25 mM HEPES KOH (pH 7.4), 5% glycerol (w/v), 0.1 mM EDTA, 180 mM NaCl, and 0.01% NP-40 (v/v)]. The column was then washed with three column volumes of Buffer A and eluted with a linear gradient of Buffer B (Buffer A containing 1000 mM NaCl). RFC-containing fractions were pooled and dialyzed overnight into Buffer C Buffer [50 mM potassium phosphate (pH 7.6), 100 mM NaCl, 5% glycerol (w/v), and 0.01% NP-40 (v/v)] at 4 ºC. Samples were loaded onto a pre-equilibrated Bio-Scale TM Mini CHT Type II 40 μM Cartridge (Bio-Rad, CA, USA) and washed with 2.5 column volumes of Buffer C. RFC was eluted with a step-wise gradient with Buffer D [500 mM potassium phosphate (pH 7.65), 100 mM NaCl, 5% glycerol (w/v), and 0.01% NP-40 (v/v)]. RFC containing fractions were then pooled and purified using a Superose6 10/300 GL (Cytiva, MA, USA) with [25 mM HEPES KOH (pH 7.4), 300mM NaCl, 2 mM DTT, 15% glycerol (w/v), 0.01% NP-40,]. RFC was concentrated in gel filtration buffer to a final concentration of ∼8 mg/ml.

#### FEN1

FEN1 was purified as described previously (29). Briefly, FEN1 was expressed in Rosetta cells containing pET28b/hFEN1-6xHis that were grown in LB supplemented with 30 µg/ml kanamycin at 37 ºC. 150 mL cultures were grown until an OD of 0.4-0.5 and then induced with 1 mM IPTG for 3 hours at 37 ºC. Cells were pelleted at 4000 x g and resuspended in Ni Lysis Buffer [50 mM NaH_2_PO_4_, 5 mM Tris (pH 8.0), 300 mM NaCl, 10 mM imidazole, 1 mM PMSF, 0.1% NP-40, and mini EDTA-free Protease Inhibitors] and sonicated. Lysates were centrifuged at 14000 x g for 10 minutes and the soluble protein was incubated with cobalt resin (Goldbio) that was pre-equilibrated with lysis buffer for 1 hour. Resin was washed with 5 column volumes of Wash buffer [50 mM NaH2PO4, 5 mM Tris (pH 8.0), 300 mM NaCl, 20 mM imidazole, 1 mM PMSF, 0.1% NP-40]. FEN1 was eluted with Ni Elution Buffer (50 mM NaH_2_PO_4_, 5 mM Tris (pH 8.0), 300 mM NaCl, 300 mM imidazole, 1 mM PMSF, 0.1% NP-40). FEN1 was then dialyzed overnight at 4 ºC in storage buffer (20 mM Tris (pH 8.0), 150 mM NaCl, 5 mM BME) and concentrated to 1 mg/ml.

### Crystallization and structural determination

PCNA-C148S was crystallized by the hanging-drop method. For crystallization, 1 µl of protein (15 mg/ml) was mixed with an equal volume of well-solution (175 mM Magnesium acetate and 20% PEG 3350). Crystals appeared after 3-7 days at room temperature. Crystals were briefly swiped through Paratone N cryo-protectant and frozen at 100K in a cryostream. Crystallographic diffraction data was collected on a Rigaku system with a Saturn 944 CCD detector. Indexing, integration, and scaling were performed with HKL3000, and the structure was solved via molecular replacement using PHASER with the partner binding region (residues 116:133) of PCNA-S228I deleted as a search model (PDB: 5E0T) (9,30). Refinement and model building was carried out using phenix.refine (31) and Coot (32). Model statistics are as defined by phenix.refine and summarized in Supplementary Table 3.

### FEN1 *in vitro* pull-downs

FEN1 was expressed and coupled to cobalt resin as described above. After FEN1 coupling, the resin was washed with 5 × 5 column volumes of Nickel Wash Buffer. The FEN1-coupled resin was then incubated for 1 hour with 5 µM PCNA-WT, C148S, or S228I at 4 ºC. The resin was then washed again with 5 × 5 column volumes of Nickel Wash Buffer to remove excess PCNA. PCNA and FEN1 were then eluted with 0.5 column volumes with Nickel Elution Buffer. Samples were separated on a 12% SDS-PAGE gel and stained with Coomassie. Densitometry was conducted in ImageJ.

### Isothermal titration calorimetry (ITC)

ITC conditions were similar to those previously used to determine peptide binding thermodynamics to PCNA-WT and S228I (9). All peptides were synthesized by 21^st^ Century Biochemicals and their sequences included: p21^CIP^ [Ac-KRRQTSMTDFYHSKRRLIFS-amide (350 µM)], FEN1 [STQGRLDDFFKVTGSL-OH (800 µM)], pol δ p66 [KANRQVSITGFFQRK (550 µM)] and RNaseH2B [DKSGMKSID TFFGVKNKKKIGKV-OH (750 µM)]. A typical reaction contained 33-38 µM individual PCNA (*i*.*e*., not trimeric rings). p21^CIP^-contraining reactions were conducted at 30 ºC while pol δ p66, FEN1 and RnaseH2B were performed at 25 ºC. The heat of dilution for reactions was subtracted by the average of the last 4-5 injection points.

### Thermal shift assay

We assessed intrinsic tryptophan fluorescence at increasing temperature conditions to obtain a relative T_m_. PCNA was diluted to various conditions (200 nM or 1 µM) in either PCNA Gel Filtration Buffer prepared using with 25 mM Tris-HCl (pH 7.5) or HEPES (pH 7.5). Tryptophan fluorescence was measured using a Fluoromax 4 Spectrofluorometer (Horiba Scientific, NJ, USA). Typical parameters were excitation: 280 nm (slit width 2nm) and emission: 325 nm (slit width 5 nm), integration time (0.1s) temperature range 25 to 60 or 70 ºC; equilibration times of 5 minutes at each temperature. Fluorescence values were normalized with Y= F -F_U_/ F_N_-F_U_ where F is the raw fluorescence signal F_U_ and F_N_ are the theoretically calculated fluorescence signals at each temperature from the unfolded and native state respectively. F_N_ and F_U_ values were determined from the linear relationship of the first and last 5 calculated values respectively. The melting point and thermodynamic values were determined from a Boltzmann sigmoidal curve. To assess the kinetics of decay, PCNA was incubated at a constant temperature for 24 hours and monitored every 2 minutes. Samples were normalized to the first time point and fit with a two-phase decay.

### Native gel assay

The stability of the PCNA trimer was determined by using a native gel assay (33). All PCNA variants were incubated for 24 hours at either 4 ºC, 37 ºC, or 42 ºC. Various concentrations of each variant were then loaded on a 4-20% gradient PAGE gel (Biorad, 4561096). Samples were run at 4 ºC in a Native Gel Buffer (192 mM glycine and 25 mM Tris base) for 2.5 hours at 100 mV. Gels were then stained with Coomassie.

### Equilibrium unfolding experiments

The stability of PCNA-WT and C148S was assessed by Circular Dichroism (CD) in the presence of incremental Guanidinium Hydrochloride [Gdm-HCl] concentrations. PCNA was dialyzed overnight into at 4 ºC in 150 mM KPO_4_ buffer (pH 7.0) with 1 mM TCEP. Samples (1µM) were then titrated with different Gdm-HCl concentrations (0-6 M) in 150 mM KPO_4_ buffer (pH 7.0) with 1 mM TCEP buffer and allowed to equilibrate for five days. CD signals were collected using a Jasco-810 spectropolarimeter (Jasco, Inc) equipped with temperature control system. Equilibrium unfolding was monitored from 215-260 nm were collected for each sample in a 2 nm cuvette at 25ºC with 25 nm bandwidth. CD signals were plotted at 218 nm versus [Gdm•HCl]. Data were fit with Savuka (34) using a two-state mode (35). The tertiary structure was measured by monitoring intrinsic tryptophan fluorescence using the same samples [excitation: 295 nm (bandpass 2 nm); emission: 325 nm (bandpass 5 nm)].

### ATPase activity assay

DNA-mediated ATPase activity of RFC was measured at room temperature using a previously published coupled-enzyme ATPase assay (11,36). Reactions mixtures containing 1 µM PCNA and 150 nM RFC were incubated with a master mix [3U/mL Pyruvate kinase, 3 U/mL Lactate dehydrogenase, 1 mM ATP 670 nM Phosphoenol pyruvate, 170 nM NADH, 50 mM Tris (pH 7.5), 5 mM MgCl2, and 400 mM Potassium glutamate] and 1 µM annealed oligos. PCNA concentrations were titrated from 0-600 nM for all affinity studies. For all the half-life experiments, PCNA (1 µM) was incubated at the designated temperature for either 1 or 24 hours prior to the experiment and diluted to a final concentration of 150 nM. All reactions were done at room temperature with reagents equilibrated for at least 10 minutes. Absorbance was measured in 96-well plates using an excitation filter at 355 nm and a bandpass of 40 nm. Oligos used in the reactions were 5’ -TTTTTTTTTTTATGTACTCGTAGTGTCTGC-3 and 5’ -GCAGACACTACGAGTACATA–3.

### Preparation of bead-based DNA template

We determined the kinetics of PCNA loading by a bead-based PCNA loading assay. ssM13 DNA (7 kbp; New England BioLabs) was coupled to streptavidin beads (ThermoFisher Dynabeads kilobaseBINDER Kit, 60101) overnight at room temperature using biotinylated oligos following manufacturer’ s recommendations. Beads were resuspended in autoclaved water and coupling efficiency was determined by PCR. Loading reaction occurred in a 1X Loading Buffer (50 mM HEPES, 4% glycerol, 0.01% NP-40, 250 mM Potassium Glutamate, 5 mM MgCl and 0.5 mM TCEP) with ∼50 ng of ssM13, 1 µM PCNA, 150 nM RFC, and 2 mM of various nucleotides. For elevated temperature experiments, PCNA was pre-incubated at 42 ºC for 24 hours prior to loading. Loading reactions were conducted at room temperature for times indicated and quenched by adding 25 mM EDTA and chilling samples on ice. Following two washes with Loading buffer, we eluted all bound protein with Laemmli buffer. PCNA was detected by Western Blot using an anti-PCNA antibody (Abcam, ab29) and quantified by ImageJ (37).

### Whole-cell and cell fractionation Western blots

Cells were grown till 70-80% confluency in 10 or 25 cm dishes, as described above. Cells were harvested and washed with 1X PBS and stored at -80 ºC until lysis. Whole-cell extracts were prepared by lysing cells in RIPA buffer (Cold Spring Harbor protocol) for 30 minutes with periodic vortexing. Cytosolic and nuclear extracts were prepared by using NE-PER™ Nuclear and Cytoplasmic Extraction Reagents (ThermoFisher, 78833) as described by the manufacturer. Chromatin fractions were prepared by sonicating the nuclear pellet in RIPA buffer for 15 minutes at 4 ºC. Proteins were detected using the following antibodies: PCNA (Abcam, ab29), Tubulin (Cell Signaling, 2146S), and H2B (Cell Signaling, D2H6). Protein levels were determined in three biological replicates by using ImageJ and compared expression HDCs.

### Protein half-life experiments

To determine the half-life of each PCNA variant, cycloheximide pulse-chase experiments were performed. Cells were incubated with either DMSO or 500 µM of cycloheximide (MP Biomedicals) at 37 ºC, 5% CO_2_ for 24 hours. Following lysis with RIPA buffer, we assessed PCNA levels by Western blot (Abcam, ab29) and measured by densitometry, as above. Values were normalized to each loading control and then normalized against the DMSO vehicle. Results represent the average of three biological replicates.

**Table 1:**
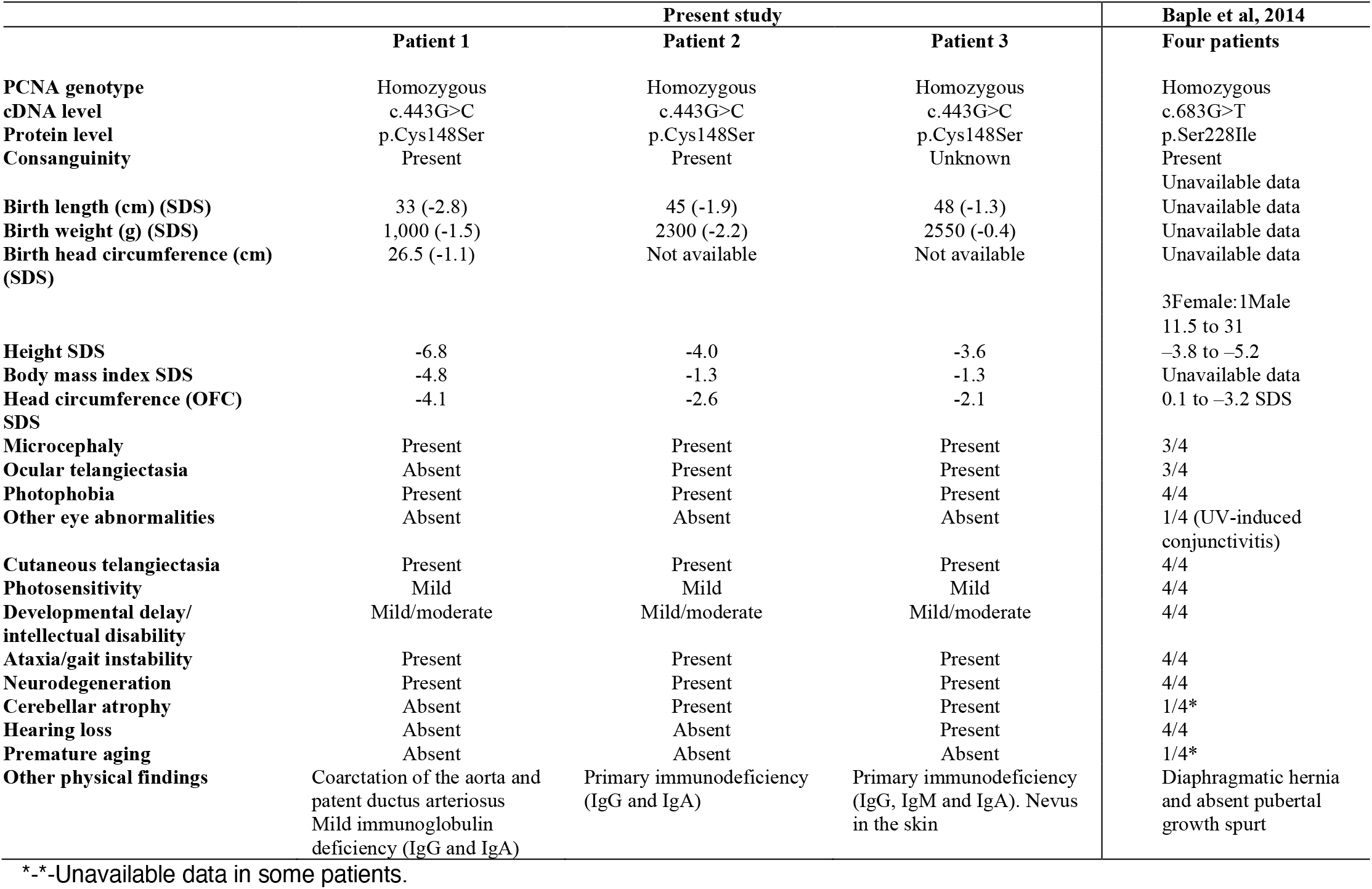
Clinical features of the patients identified with pathogenic variants in *PCNA* gene associated with PARD.

### Flow cytometric γH2AX signaling

Exponential growing cells were plated the day before differential temperature treatment and incubated at 37 ºC. Cells were separated to different temperature (37 or 42 ºC) for 24 hours. At the indicated time points, cells were collected as previously described (72). Briefly, cells were fixed on ice with 1% formaldehyde and storage at -20 ºC in 70% ethanol. Cells were permeabilized and blocked with BSA-T buffer (0.2% Triton X-100 [Sigma–Aldrich] and 1% BSA in PBS). Cells were then incubated with 1/5000 anti-γH2AX (05-636 Millipore) overnight at 4 ºC. Samples were incubated overnight at 4 ºC with 1/200 FITC anti-mouse secondary antibody (Sigma– Aldrich). Cells were then stained with propidium iodide solution (PI -20 μg/ml, 200 μg/ml RNase A -Invitrogen, Life Technologies and 0.1% Triton X-100) in the dark and at room temperature for 30 min. Cells were processed on a BD AccuriTM C6 (BD) type cytometer and analyzed using the BD CSamplerTM Analysis software. More than 10,000 events were obtained for the analysis of each sample.

## RESULTS

### Clinical description of the patients 1, 2 and 3

Patient history removed for medRxiv standards.

Patient history removed for medRxiv standards.

### Whole exome sequencing of patient samples reveals the presence of a homozygous PCNA mutation

To identify the genetic cause underlying the observed disorders, we performed whole-exome sequencing (WES) on all three patients. In all cases, the average coverage of the target exonic regions was >105x with a coverage range of >96%. We filtered sequence variants by focusing on rare deleterious homozygous variants across all three patients, identifying 6 to 15 missense mutations in each patient (Supplemental Table 1).

We found that the only potentially deleterious variant shared amongst all three patients is a homozygous transversion mutation within exon five of the *PCNA* gene, (variant ref ID: NM_002592.2: c.443G>C). The resulting mutation causes a cysteine to serine alteration at position 148 of the PCNA protein ((p.C148S), NCBI dbSNP ID: rs1274412848) (38). We confirmed that this mutation is homozygous in all three patients using Sanger sequencing (Figure 2B). We also determine the genetic status of healthy family members by Sanger sequencing and found that none of them were homozygous for PCNA-C148S. This allele is very rare in the general population, as it is only found at a very low frequency in the gnomAD database (Minor allele frequency of 0.00002897 in the neighboring population). Additionally, our analysis revealed all three patients share common polymorphisms within ∼250 kb (Chr20:4,972,112-5,221,841) around the *PCNA* gene (Chr20: 5,098,255). While this observation suggests that the three patients share a common ancestor, all three reside in distinct regions of the country. The fact that they have no known shared family history (Supplementary Figure 1) led us to conclude that they are distantly related relatives. The C148 residue is highly conserved across all metazoan species (Figure 2G, Supplemental Figure 1) and the C148S variant is predicted to be pathogenic (Supplemental Table 2). Taken together, our results collectively suggest that the underlying cause of all three patients’ disease is due to the presence of the C148S substitution in PCNA.

**Figure 2:**
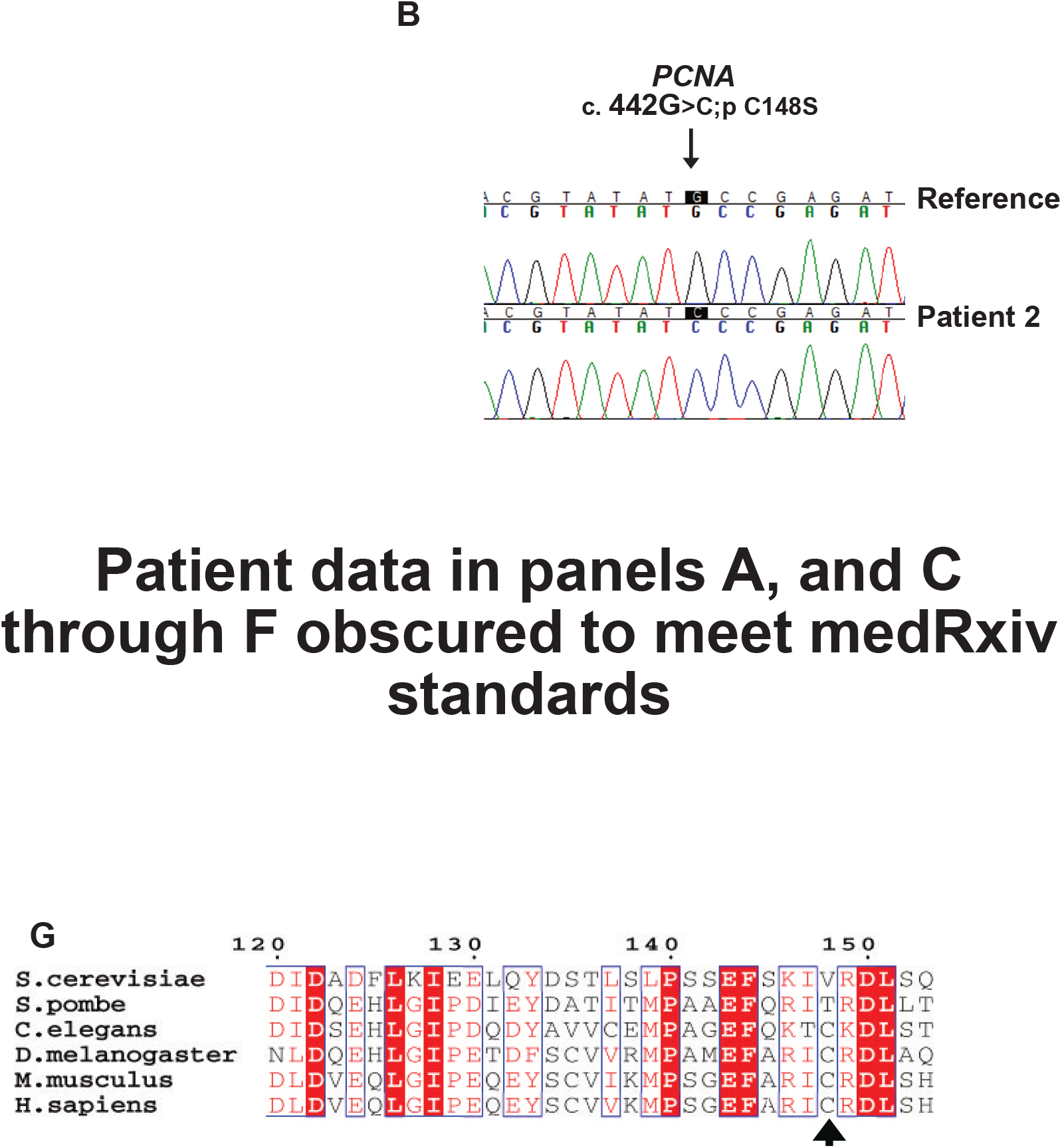
A novel Ataxia Telangiectasia-like syndrome associated with an alteration in a highly conserved region of PCNA. **(A)** Pedigree tree of the three patients. The known genotype of each individual confirmed by Sanger sequencing is below each symbol (“Wild” is the reference allele and “Mut” is the altered allele). **(B)** Sanger sequencing confirming the point mutation (c. 442G.C>; pC148S) in the PCNA gene. (C-F) clinical presentation of patients with the C148S allele. Patients present with **(C)** mild facial dysmorphisms: broad forehead, low-set posteriorly rotated ears, broad nasal bridge, thin upper lip, and cutaneous telangiectasia located on face **(D)** ocular telangiectasia, **(E)** systemic cutaneous telangiectasia **(F)** neurodegeneration. See Supplemental Text for the complete clinical presentation of each patient. **(G)** Protein sequence alignment of PCNA from several species. A cysteine at position 148 is conserved across multi-cellular eukaryotes.

### Patient-derived-C148S cells have increased sensitivity to double-stranded DNA damaging agents

We next investigated whether the C148S variant causes a defect on cell viability after DNA damage. Because cells harboring the S228I variant have increased sensitivity to UV-induced DNA damage (8), we wondered if PCNA-C148S cells exhibit a similar phenotype. To address this question, we measured cell viability of patient-derived fibroblasts with increasing dosage of UV-irradiation (Figure 3A). As a reference, we used two different healthy donor cells (HDCs) as negative controls and fibroblasts from Cockayne syndrome patients (polη deficient) as positive controls. We found that unlike PCNA-S228I, C148S fibroblasts were as resistant to UV irradiation as HDCs (Figure 3A).

**Figure 3:**
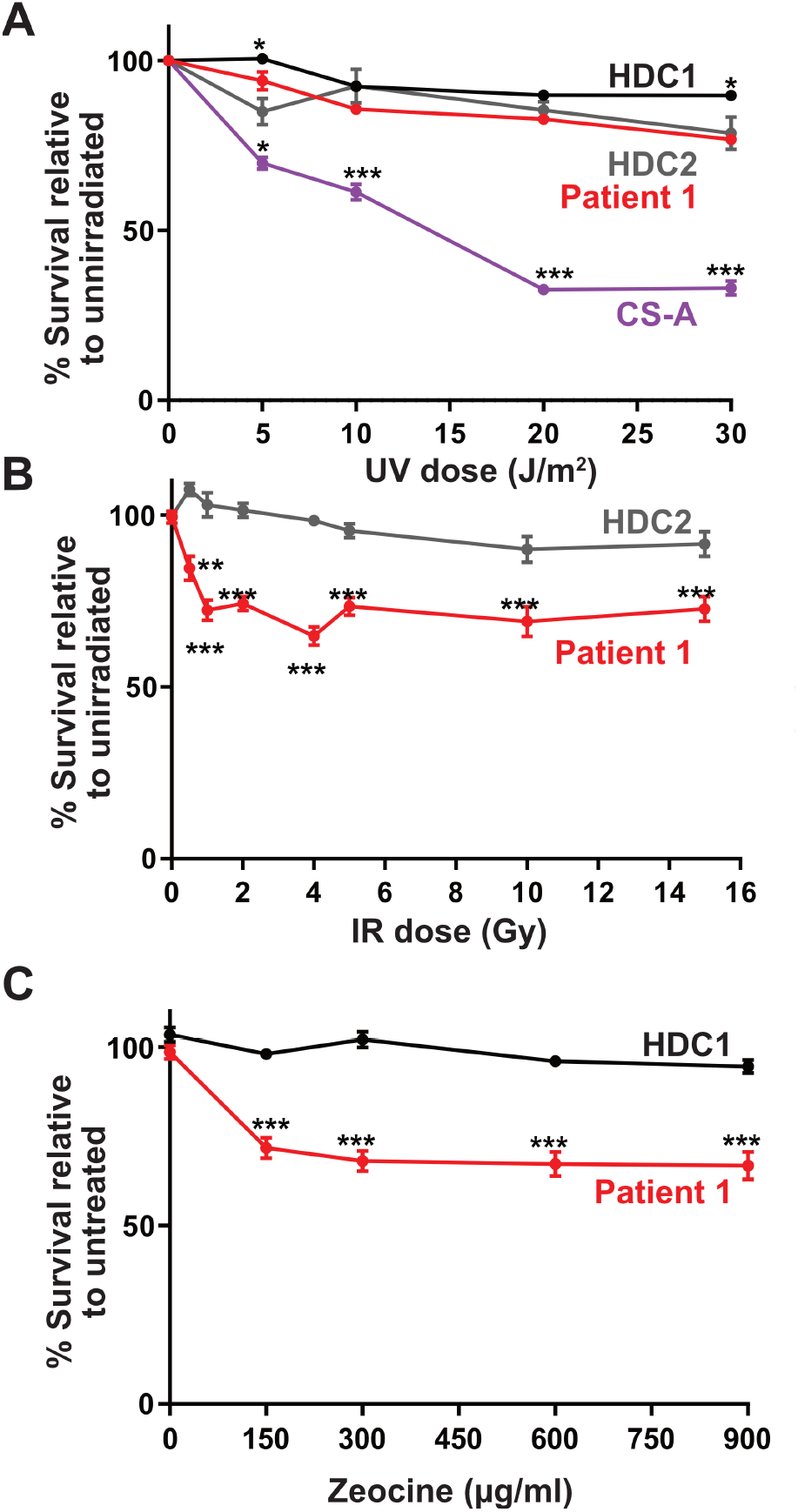
Cells harboring the C148S allele are sensitive to dsDNA break damage. **(A)** PCNA-C148S are not especially sensitive to UV-induced DNA damage. Cells were treated with increasing dosages of UV and their cell survival was determined by XTT assay (n=6). **(B-C)** PCNA-C148S cells are very sensitive to dsDNA damage agents. Cells were treated with either increasing dosages of IR or zeocine and then measure for cell viability (n=6). (p-values from multiple t-tests *<0.05, **<0.01, ***<0.001).

Because cells defective in DNA replication and repair machinery are often sensitive to dsDNA damage agents, (39,40), we next investigated whether C148S fibroblasts were sensitive to increasing dosages of ionizing radiation (IR, Figure 3B). We found that patient-derived C148S cells were more sensitive to IR-induced DNA damage compared to HDCs. We confirmed our results by pharmacologically inducing dsDNA breaks with zeocine, and again observed an increased sensitivity in C148S cells compared to HDCs (Figure 3C). Our combined results suggest that the C148S mutation impairs dsDNA break repair, which manifests as reduced viability, but does not increase sensitivity to UV-induced damage. This is a significant distinction between patient-derived C148S and S228I fibroblasts, which we decided to investigate biochemically.

### No major conformational change in PCNA-C148S

We tested whether the C148S substitution affects the overall structure of PCNA, as was observed with S228I (10). We determined the crystal structure of PCNA-C148S to 3.1-Å resolution (Figure 4A, Supplemental Table 3). We found that overall the structure of PCNA-C148S is similar to that of PCNA-WT with an RMSD value of 0.85 Å, indicating no significant global changes to the structure. Furthermore, the conformation of residues surrounding C148S does not differ significantly from PCNA-WT. Although the S228I variant has a deformed partner binding cleft (9,10), this region is not obviously altered in PCNA-C148S. It is important to note that a portion of the binding cleft exhibits weak density in our PCNA-C148S structure. This challenge prevented precise modeling of the entire partner binding cleft. Despite this issue, there is unambiguous density for Y133, a residue that dictates the conformation of the partner binding cleft (9,10). Because Y133 is in the same conformation as seen in PCNA-WT, we conclude that the partner binding cleft is unchanged in PCNA-C148S (Figure 4A).

**Figure 4:**
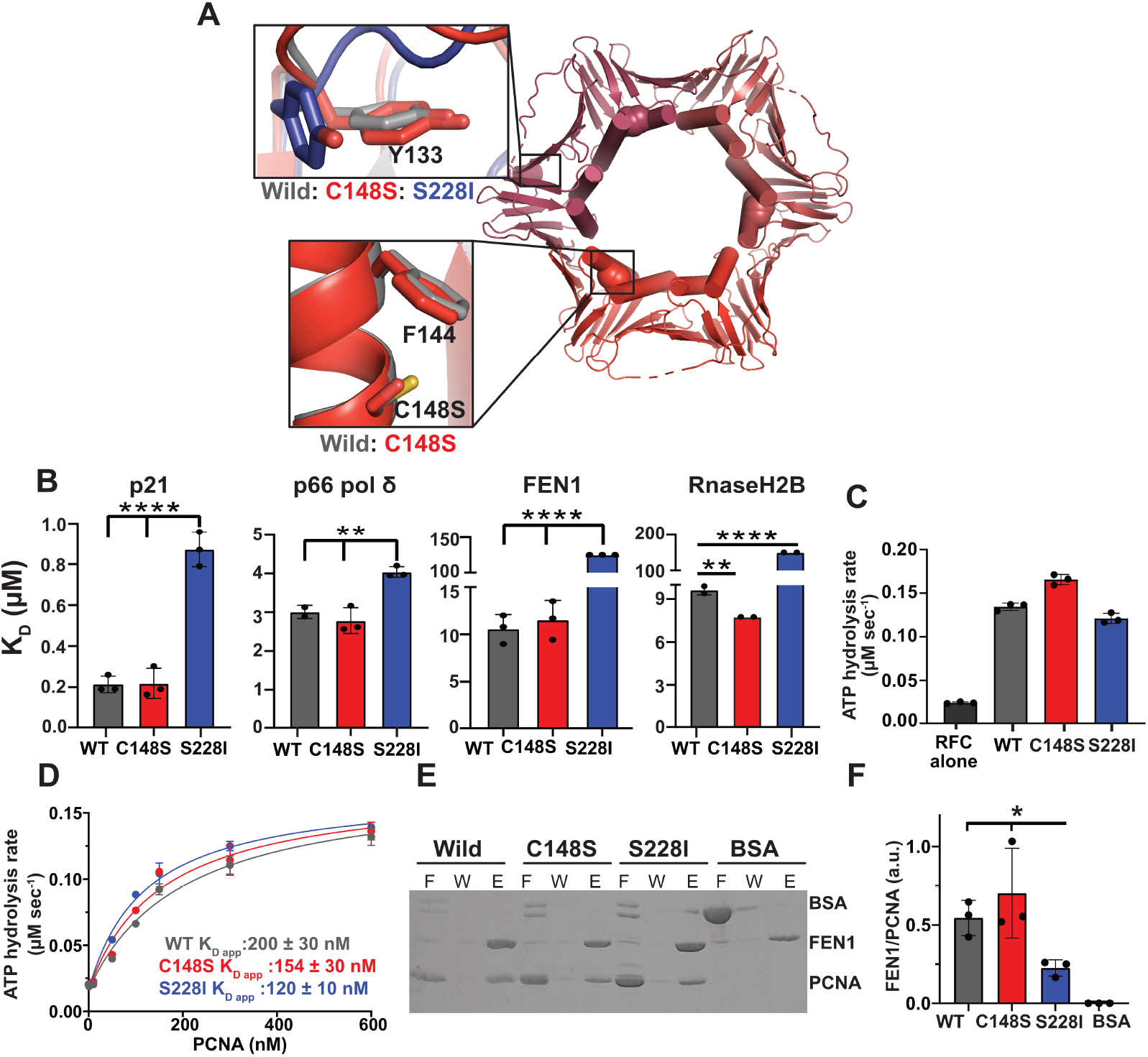
PCNA-C148S is structurally similar to and binds to partners like wild-type. **(A)** A 3.1-Å resolution crystal structure of PCNA-C148S (Red shades; S148 highlighted in each subunit as spheres) shows no structural changes in the globular portion of the protein. Weak density was observed for the partner binding region of PCNA-C148S. The upper-left box shows that Y133 is similar to WT-PCNA (Gray, PDB: 1VYM), which was rotated ∼90º in PCNA-S228I (Blue, PDB: 5E0T, Upper-right box). The cysteine to serine substitution does not cause structural changes to itself or surrounding residues (lower-left box). **(B)** PCNA-C148S binds to four different partners peptides like wild-type as determined by Isothermal Titration Calorimetry (ITC; n= 2 or 3; p-values from one-way ANOVA with multiple comparison against PCNA-WT; **<0.005, ****<0.0001). As a point of comparison, PCNA-S228I measurements are from Duffy *et al*. 2016. See Supplemental Figure 2 for raw ITC titration curves. For FEN1 and RNasH2B, values are the theoretical K_D_ based on the detection limit. **(C-D)** Both PARD variants stimulate RFC clamp loader ATP hydrolysis and bind like PCNA-WT (n=3). WT and PCNA-S228I are rates from Gaubitz *et al*. 22020 and were collected on the same day as PCNA-C148S. **(E-F)** PCNA-C148S, but not PCNA-S228I, binds to full-length FEN1 by *in vitro* his-pulldown (F= flow through, W= wash, E= elution; n=3; p-value from t-test * <0.05).

### The C148S substitution does not impair partner binding

Because PCNA-S228I is defective for binding to numerous partner proteins (9, 10), we sought to investigate if a similar defect exists with PCNA-C148S. We focused our efforts on four different PCNA partners: p21^CIP1^, DNA polymerase δ (pol δ), FEN1, and RNaseH2B, as these proteins participate in various DNA replication and/or damage response pathways, and their affinity for PCNA-WT and/or S228I were previously reported (9,10,41). We measured partner PIP-box peptide affinity by isothermal titration calorimetry (ITC). We found no significant difference in binding affinity between PCNA-WT and C148S (Figure 4B; Supplemental Figure 2 & Table 4), except for a subtle increase in affinity for RNaseH2B. These results are in contrast to PCNA-S228I, which binds pol δ, FEN1 and RnaseH2B poorly (9,10), indicating that the two associated PARD variants do not share the same partner binding properties.

To see if these differences in binding extend to full-length partners, we measured binding to two full-length partners (*i*.*e*., RFC and FEN1) were used instead of PIP-box peptides. PCNA is known to stimulate the ATPase activity of RFC (3,11,42), and we therefore leveraged this feature to assess RFC binding to PCNA. PCNA-WT, C148S and S228I stimulated the ATPase activity of RFC to the same order of magnitude (Figure 4C). We then determined affinity of PCNA for RFC by measuring ATPase activity as a function of PCNA concentration (Figure 4D), as previously reported for PCNA-S228I (11). We found that PCNA-WT, C148S and S228I all have similar affinity for RFC (K_d, app_ ∼ 150 nM, 200 nM, and 120 nM, respectively). We next assessed the binding affinity between FEN1 and PCNA through a pull-down experiment using His-tagged full-length FEN1. As expected, PCNA-S228I binds FEN1 poorly (9), while both WT-PCNA and C148S bind similar affinities for FEN1 (Figure 4E-F). Our combined structural and affinity binding studies indicate that, in contrast to PCNA-S228I, PCNA-C148S does not exhibit the defect in partner binding that was thought to drive PARD.

### PARD mutations impair PCNA stability

From a structural point, the C148 residue is buried in a hydrophobic pocket near the interface between subunits and participates in a π-sulfur interaction with F144 (Figure 4A). These types of interactions have been shown to confer substantial protein stability (43). Thus, we hypothesized that the C148S substitution disrupted protein stability. To investigate this hypothesis, we measured thermostability of PCNA using a thermal shift assay, with tertiary structure monitored by PCNA’ s intrinsic tryptophan fluorescence. We found that all three PCNA variants display unfolding curves consistent with a two-state unfolding mechanism (Figure 5; Supplemental Figure 3 & Table 5). Both PCNA-C148S and S228I have dramatically reduced thermal stability compared to PCNA-WT (Wild-type T_m_: 52.0 ± 0.03 ºC, PCNA-C148S T_m_: 42.0 ± 0.06 ºC, and PCNA-S228I T_m_: 44.0 ± 0.1 ºC). Based on our data, we estimate that ∼17-22% of PCNA-C148S and -S228I are in the unfolded state at normal physiological temperature. To quantify the energetic consequences of the C148S substitution on PCNA stability, we conducted equilibrium unfolding experiments. We incubated PCNA-WT and C148S with different concentrations of the denaturant agent guanidinium hydrochloride (Gdm-HCl) and monitored secondary structure by circular dichroism (CD) and tertiary structure by tryptophan fluorescence. Both readouts exhibit a steep transition at identical Gdm-HCl concentrations, indicating a single, cooperative transition between the folded and unfolded state (Figure 5B). In addition, our results showed that PCNA-C148S unfolds at a lower concentration of Gdm-HCl than wild-type (*i*.*e*. ∼0.5 M), verifying a stability defect through independent approaches. Our results suggest that the C148S substitution causes a ∼0.6 kcal/mol loss in folding free energy; however, the precision of this estimate is not entirely clear due to the noisy baselines in the PCNA-C148S data. Collectively, our results indicate that PCNA-C148S and -S228I are significantly less stable than wild-type and that a fraction of each variant is unfolded at physiological temperatures.

**Figure 5:**
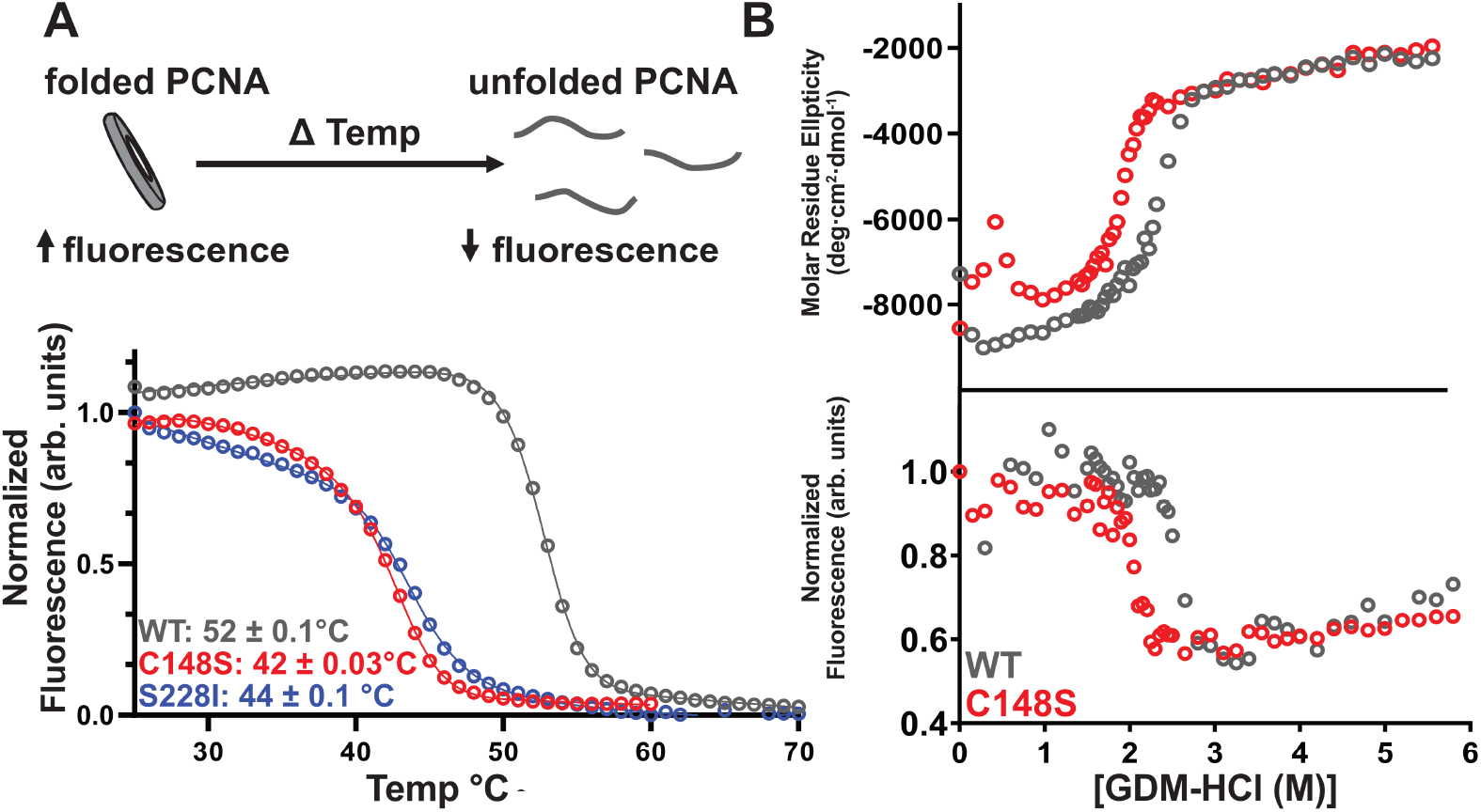
PARD variants are less stable than wild-type. **(A)** PCNA tryptophan fluorescence decreases as PCNA unfolds (top cartoon). Below is a representative thermal shift curve (1 µM PCNA). **(B)** PCNA-C148S is less stable than wild-type in the presence of Guanidinium-HCl (Gdm-HCl). Top graph is the normalized circular dichroism (CD) signal that measures secondary structure. Bottom graph is normalized tryptophan fluorescence, which is a proxy for tertiary structure. The free energy of each variant was determined from the CD signal (wild-type: Δ G= 8.20 ± 0.05 kcal/mol; C148S: Δ G= 7.6 ± 0.6 kcal/mol)

### PARD variants are quickly inactivated at elevated temperature

Noting the stability defect of the PARD associated variants, we next asked if the substitutions compromised the formation of functional trimers. We measured PCNA oligomerization after incubation at 4ºC, 37 ºC, and 42 ºC using a native polyacrylamide gel assay (Figure 6A, (33,44)). As a point of comparison, we used PCNA-D150E, a substitution that is known to disrupt trimerization, and lead to monomerization (45,46). We found that PCNA-WT, -C148S, -S228I all migrated as trimers after incubation at 4 ºC, while D150E migrated as an expected monomer. In contrast, pre-incubation at 42 ºC led to the formation of higher molecular bands indictive of aggregation for all PCNA variants except PCNA-WT, which primary behaved as a trimer. We did observe that a small fraction of PCNA-S228I persisted as a trimer, which is consistent with its slightly higher melting point. This heat-induced aggregation also occurred at 37 ºC for both PARD-associated variants, but to a lesser extent than at 42 ºC, consistent with our tryptophan fluorescence data (Figure 5A & Supplemental Figure 4A).

**Figure 6:**
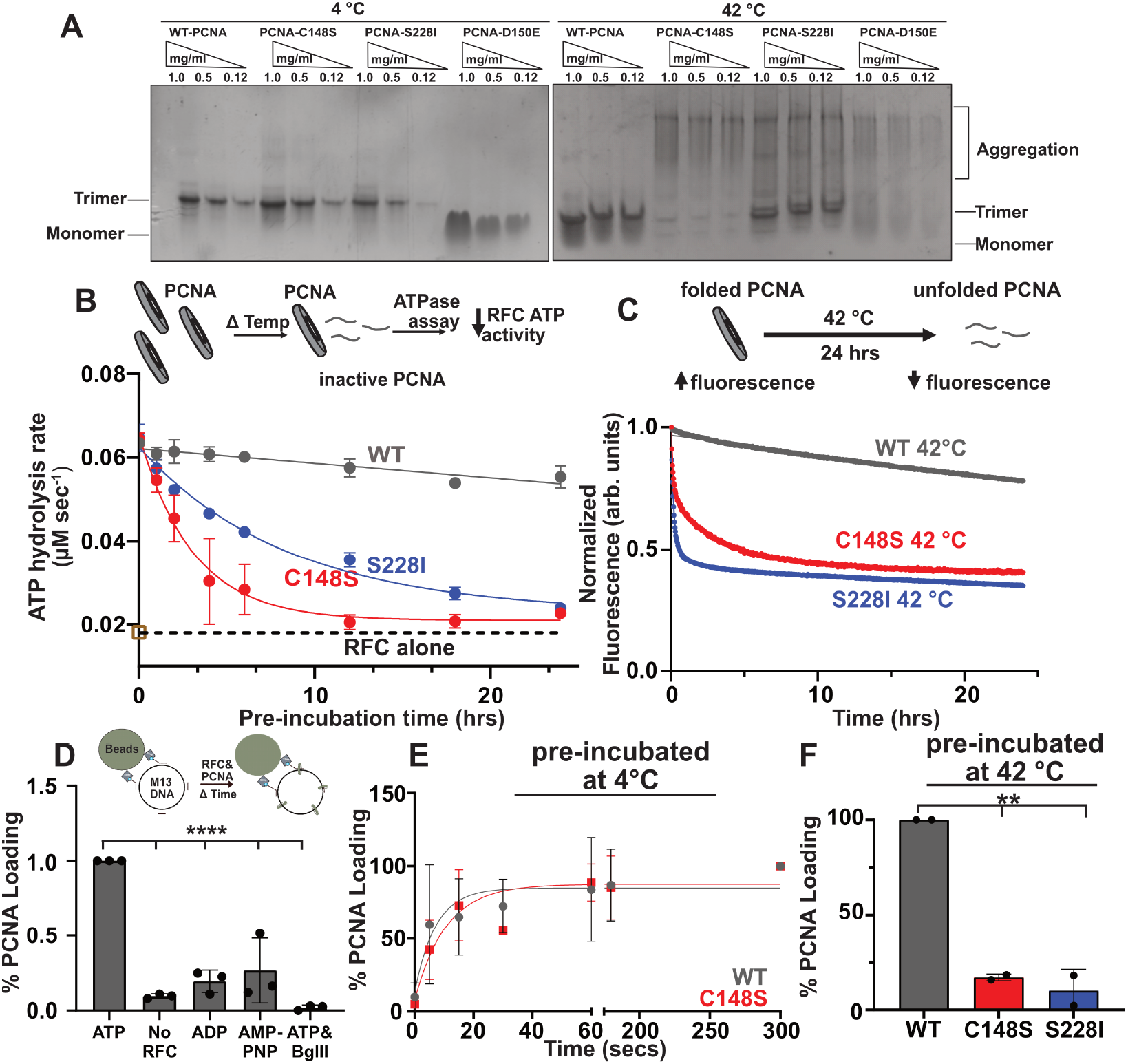
PARD variants are inactivated by heat. **(A)** Temperature induced aggregation of both PARD variants. All PCNA variants were incubated at either 4 ºC and 42 ºC for 24 hours and separated on a 4-20% Native PAGE Gel stained with Coomassie blue. **(B)** Both PARD variants are rapidly heat-inactivated and fail to stimulate RFC. Heat inactivation over time causes denatured PCNA, which fails to stimulate RFC (top cartoon). Below are the fitted curves, where the dotted line is the intrinsic activity of RFC alone (n=3; see Supplemental Figure 4). **(C)** Both PARD variants have an increased rate of decay *in vitro*. PCNA signal decreases over time when held at a constant 42 ºC for 24 hours (top cartoon). Below are the fluorescence curves for each PARD variant. **(D)** Bead-based PCNA loading assay directly measures PCNA loading. M13 DNA is coupled to magnetic beads *via* streptavidin-biotin interactions. DNA-beads are incubated with PCNA and RFC over time. Excess PCNA and RFC are washed away and the amount of PCNA on the beads is measured via Western blot (top cartoon). Western blot signal of PCNA detected in the presence of different conditions is shown below (n=3, p-value from one-way ANOVA and multiple comparison ****<0.0001) **(E)** When preincubated at 4 ºC PCNA-C148S loads onto DNA like wild-type. **(F)** Both PARD variants fail to load onto DNA after 24-hour preincubation at 42 ºC (n=2, p-value from one-way ANOVA and multiple comparison **<0.005). See supplemental for representative images of PCNA loading assays **(5D-F)**.

We next investigated how each PARD substitution impacts the functional half-life of PCNA at various temperatures. Because RFC’ s activity depends on functional-trimeric PCNA, we used its ATPase activity to measure levels of functional PCNA (Figure 6B). We pre-incubated each variant for 1 or 24 hours at 4 ºC, 25 ºC, or 42 ºC, and then measured ATPase activity. All PCNA variants retain the ability to stimulate RFC ATPase activity when pre-incubated at 4 ºC or 25 ºC (Supplemental Figure 4B-D). In contrast, 42 ºC pre-incubation completely inactivated both PARD-associated variants, with the remaining ATPase activity reduced to baseline. We then calculated the “functional” half-life of each variant by pre-incubating at 42 ºC for various time points. Both PARD-associated variants decay ∼1000 fold faster than PCNA-WT at 42 °C (PCNA-C148S t_1/2_: 2.1 ± 0.05 hrs, PCNA-S228I t_1/2_: 5.9 ± 0.01 hrs, PCNA-WT t_1/2_ > 2000 hrs (Figure 6B)). This rapid decay of ATPase stimulation correlates with the kinetics of PCNA unfolding as assessed by tryptophan fluorescence at both 42 ºC and 37 ºC (Figure 6C and Supplemental Figure 4E). For example, at 42 ºC, PCNA-WT unfolds orders of magnitude more slowly than the PARD-associated variants (PCNA-C148S t_1/2_ ∼ 2.9 hrs, S228I t_1/2_ ∼ 7.3 hrs, PCNA-WT t_1/2_ ∼ 90 hrs). Thus, both PARD-associated variants are thermo-inactivated due to their faster unfolding kinetics at elevated temperatures.

Because PCNA is functional only when bound to DNA, we next asked if heat-inactivation of each PARD-associated variant impacted their ability to load onto DNA. To address this, we used a bead-based assay that allowed us to monitor the loading of PCNA onto a single-stranded DNA plasmid (Figure 6D & Supplemental Figure 5A-B). After 5 minutes, a distinct signal for PCNA was detected that is dependent on both RFC and ATP. We confirmed this assay by linearizing the DNA with a restriction enzyme (i.e. *BglII*), which allows PCNA to slide off the DNA. This eliminated the PCNA signal, indicating that PCNA is specifically loaded onto the DNA substrate. With this system, we monitored loading of PCNA that was either pre-incubated at 25 ºC or 42 ºC. We found that PCNA-WT and -C148S load onto DNA at similar rates when pre-incubated at 25 ºC (t_1/2_: 12-15 secs; Figure 6E) and to a similar extent for PCNA-S228I (Supplemental Figure 5C). In contrast, after incubation at 42 ºC, both PARD-associated variants load onto DNA at ∼10x lower levels compared to WT-PCNA (Figure 6F & Supplemental Figure 5D). Collectively, our results suggest that both PARD-associated variants function as temperature-sensitive mutants that form trimeric PCNA at low temperatures, but unfold and become inactivated at physiological temperatures and above.

### Patient-derived-C148S cells have lowered levels of PCNA

Given that both PARD-associated variants have decreased stability, we next asked how these substitutions impact total PCNA levels in patient-derived cells. To test this, we compared PCNA levels from patient-derived cells versus HDCs. We detected significantly lower PCNA levels in patient-derived cells harboring the C148S substitution compared to HDCs (Figure 7A). We next asked if these lower PCNA levels are altered in specific compartments in the cell. Therefore, we fractionated whole-cell lysates and found consistently lower levels of PCNA in all cellular compartments of patient-derived cells (Figure 7B). Most strikingly, we detected much lower chromatin-bound PCNA in PARD-derived cells than HDCs, suggesting that the C148S substitution impacts the levels of PCNA on DNA in cells.

**Figure 7:**
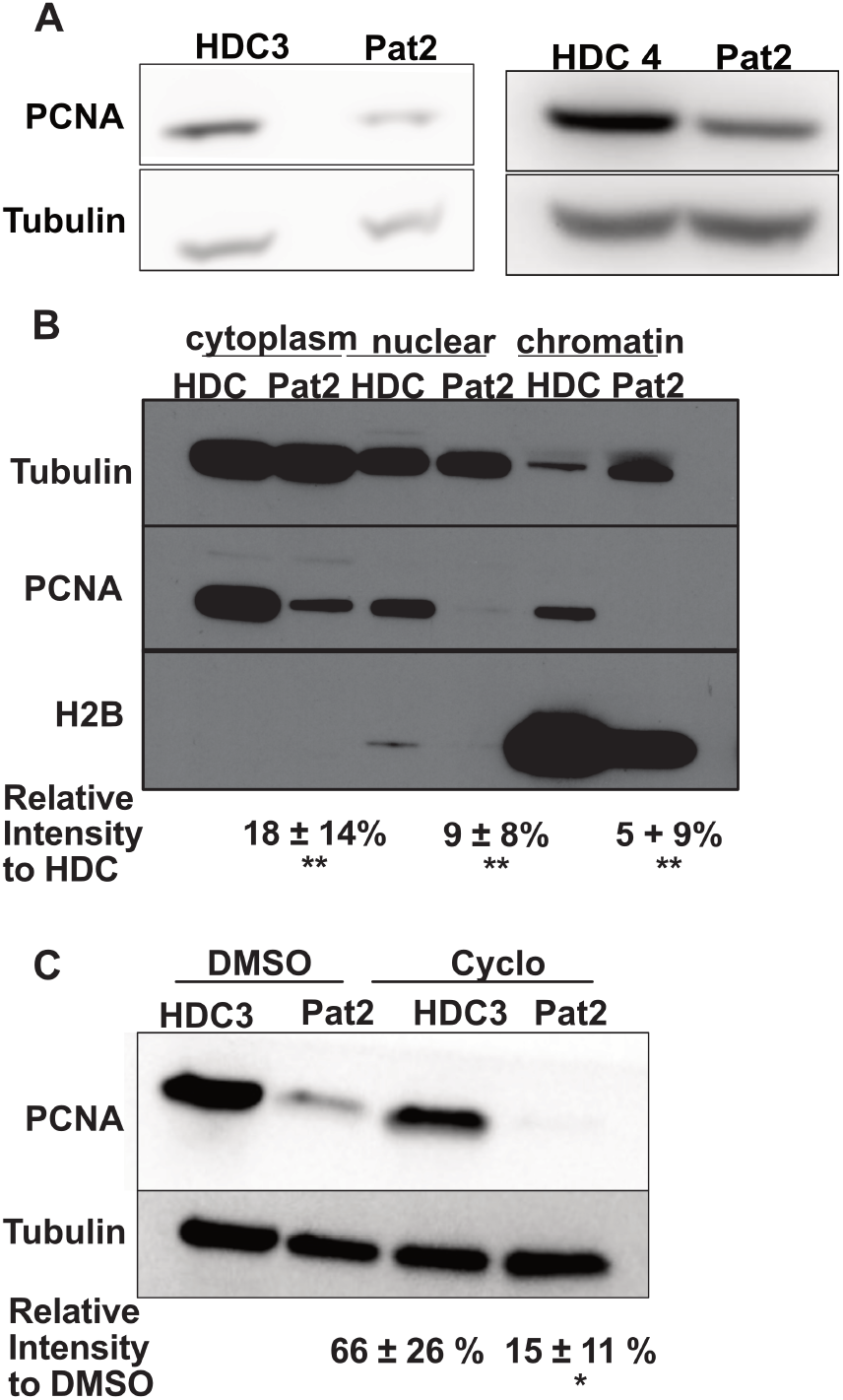
Protein levels of PCNA-C148S are lower and less stable *in cellulo*. **(A)** PCNA levels in primary fibroblasts are lower compared to HDCs. β-Tubulin was used as a loading control. **(B)** Fractioned PCNA levels of primary fibroblast show overall lower PCNA levels, especially in the nuclear and chromatin fractions. **(C)** PCNA-C148S are less stable than wild-type after cycloheximide pulse chase. All samples are normalized to the loading control β-Tubulin and to their corresponding DMSO sample for fold change differences (n=3; p-value from t-test *<0.05, **<0.001). All cells were from asynchronous populations.

We next asked if decreased PCNA-C148S stability could drive the lower levels of PCNA in patient-derived cells.Therefore, we performed cycloheximide pulse-chase experiments to monitor PCNA lifetime in cells (Figure 7C). We found after 24 hours of cycloheximide treatment, the levels of PCNA in the patient-derived cells decreased much faster than in the HDCs. Therefore, our combined molecular and cellular results indicate that PCNA-C148S is less stable than PCNA-WT, and that this defect reduces the levels of PCNA on DNA.

### Patient-derived-C148S cells exhibit defects at elevated temperatures

Noting the thermal stability defect of PCNA-C148S, we next asked if the cellular dsDNA sensitive phenotype correlated with PCNA instability. We reasoned that elevated temperatures should impact PCNA activity and therefore increase the susceptibility of PARD cells to dsDNA break-inducing agents. To test this, we compared the levels of γH2AX, a known marker of dsDNA breaks (47), at both 37 °C and 42 °C in patient-derived and control cells using flow cytometry (Figure 8A). We found that PCNA-C148S cells have a slightly higher level of γH2AX at 37 °C compared to HDCs, but at 42 °C the levels of γH2AX is much higher than in HDCs. These results suggest that the patient derived cells have higher levels of basal dsDNA breaks, which is exacerbated at higher temperatures.

**Figure 8:**
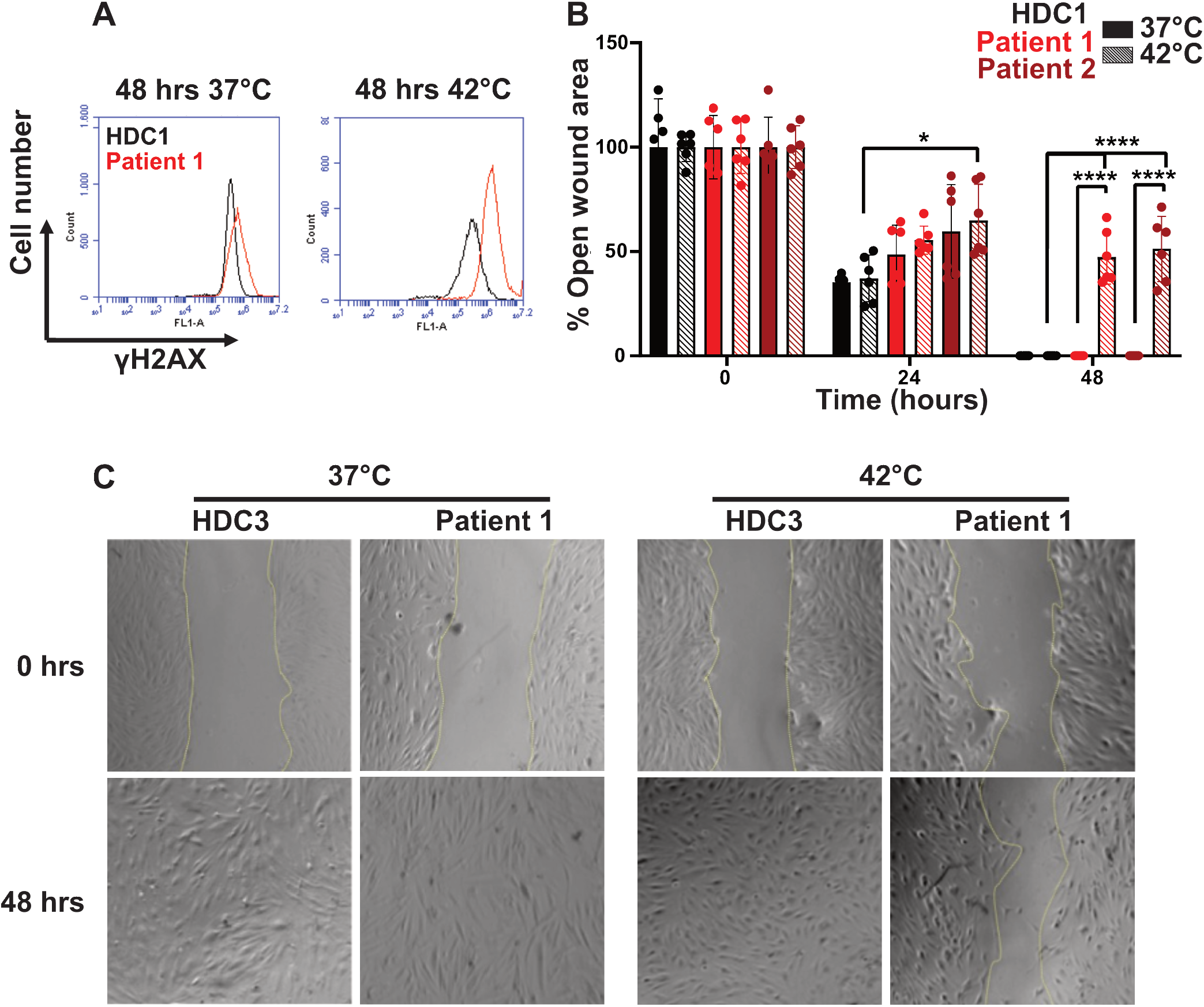
PCNA-C148S cells have a temperature-sensitivity phenotype. **(A)** Primary PCNA-C148S fibroblasts have an intrinsically higher γH2AX signal compared to HDCs, which is exacerbated at elevated temperatures. γH2AX levels were determined by flow cytometry (>10,000 events) on an untreated asynchronous population of cells. **(B)** Primary PCNA-C148S cells fail to grow like HDCs at elevated temperatures. Wound healing assays were performed with cells incubated for 48 hours at two different temperatures at time zero (n=6; p-value from t-test *<0.05; ****<0.0001). **(C)** Representative images of the wound-healing assay (see Supplemental Figure 6 for additional images).

Because γH2AX signaling can trigger cellular arrest (47), we next tested whether the observed temperature-induced increase in γH2AX expression resulted in decreased cellular growth. Therefore, to monitor the growth of each cell line, we performed a wound-healing assay at both 37 ºC and 42 ºC (Figure 8B&C and Supplemental Figure 6). Following cell adhesion, the monolayer was mechanically disrupted using a pipet tip. We then monitored “wound” healing over time by light microscopy. At 37 ºC, all cell lines were able to grow back into a single monolayer within 48 hours. In contrast, at 42 ºC, only the HDCs showed complete recovery, while the patient-derived cells harboring PCNA-C148S showed minimal growth. Therefore, these results suggest that cells harboring the C148S variant have restricted proliferation at elevated temperatures. Collectively, our findings suggest that elevated temperatures impair cell viability and provides a step towards a molecular understanding for PARD.

## DISCUSSION

### The PCNA-C148S allele is associated with PARD

Here we describe three individuals homozygous for the PCNA-C148S variant with symptoms nearly identical to the recently discovered disease PARD. Like patients with the previously reported PCNA-S228I PARD variant (8), these patients present with an atypical collection of symptoms that include: neurodegeneration, skin photosensitivity, cutaneous abnormalities, and developmental disorders. At the cellular level, both variants appear to differ from each other. Unlike PCNA-S228I, PCNA-C148S fibroblasts appear insensitive to UV-induced damage, but rather sensitive to dsDNA break-inducing agents. (The susceptibility for PCNA-S228I remains unknown (8)). We cannot completely rule out that all PCNA-C148S cells are not sensitive to UV-induced damage, as our study only focused on one cell type (fibroblasts). Regardless, our combined clinical, genetic, and experimental findings indicate that the presence of a homozygous PCNA-C148S is a novel cause of PARD.

In general, patients with either PCNA-C148S or -S228I present with symptoms similar to other DNA repair disorders. The sensitivity to sunlight and photophobia typical of PARD is also observed in both Xeroderma Pigmentosum and Cockayne Syndrome patients, although to a lesser extent (48,49). Furthermore, the ataxic gait and telangiectasia seen in PARD and Ataxia Telangiectasia patients are similar (8,50). Like these DNA repair disorders, PARD appears to have a progressive clinical component. For instance, continuous neurodegeneration likely leads to cerebellar atrophy, as these symptoms are present in the two older patients. Similarly, progressive cerebellar ataxia is observed in AT patients (51). These similarities and clinical overlap with other DNA repair disorders underscore the importance of developing genetic testing is necessary approaches to distinguish PARD from other DNA repair disorders.

Despite the central role played by PCNA in DNA replication, PARD patients present with symptoms distinct from those of other DNA replication disorders. For instance, aside from the mild immunoglobulin deficiency seen in patients expressing PCNA-C148S, PARD patients generally do not have significant immunologic dysfunction (9). This finding contrasts with the severe immunological symptoms observed in other DNA replication disorders such as Immunodeficiency 96, which results from Ligase 1 missense mutations, and Immunodeficiency 54, which is a consequence of MCM4 truncation. These individuals suffer from extreme viral and bacterial infections due to extreme leukopenia (52–54). Similar immune dysfunctions were reported in two individuals with DNA polymerase δ mutations (55). In PARD patients, the hypomorphic activity of PCNA may be sufficient to ensure normal immune function, particularly if DNA replication is not affected. However, PARD patients may have compromised immunologic functions in settings of elevated body temperatures (*e*.*g*. during gestation stages or febrile episodes) and require further surveillance. Thus, it is critical to characterize the full spectrum of symptoms that may arise from mutations in replication proteins, despite their overlapping cellular functions.

### Impact of the C148S substitution on PCNA stability

Our work provides new insight into how subtle mutations can profoundly affect PCNA stability. The C148 side-chain makes a π-sulfur interaction with the neighboring Y144 (1). In general, π-sulfur interactions provide considerable stability (∼0.5 kcal/mol), comparable to what we observed for PCNA-C148S (43). Additionally, converting a buried free cysteine to the more hydrophilic serine has been found to destabilize proteins (56– 58). Thus, two non-mutually exclusive biophysical explanations exist for the stability defect observed in in PCNA-C148S. We also observe a stability defect in PCNA-S228I, which is likely caused by the steric disruption of the partner binding cleft (9,10).

Subtle mutations that compromise trimer stability have been discovered in PCNA proteins across metazoan species (33,44,45). However, unlike most of the other reported mutations, the S228I and C148S substitutions are not directly involved in forming the interfaces between PCNA subunits. Therefore, our work demonstrates that substitutions in many regions of PCNA can potentially result in a stability defect and have harmful effects on human health. Further genetic surveillance will be necessary to reveal the full repertoire of mutations that can lead to PARD.

### PCNA-C148S provides novel insight into PARD

Our study on PCNA-C148S provides new insight into the molecular basis behind PARD. PCNA-S228I has a partner binding defect that impairs interactions with specific partners (8– 11). This does not appear to be the primary defect underlying PARD, as PCNA-C148S appears to interact with its partners with similar affinity as PCNA-WT. However, our work reveals that the only shared phenotype of between both PARD mutants is their remarkable thermostability defect (Figure 5). Furthermore, these variants appear partially impaired at physiological body temperatures (Supplemental Figure 4A), but are still permissive enough to support human life. While our findings do not negate the idea that aberrant partner binding can play a role in PARD (particularly for the S228I variant), they reveal a potential second mechanism that likely underlies this disease: a stability defect leading to lower PCNA levels on chromatin.

Maintaining proper levels of chromatin-bound PCNA is critical for genome stability. During late G1 phase, chromatin-bound PCNA levels increase in accordance with the DNA replication machinery (59). PCNA is removed from chromatin either through passive dissociation or actively by the ATAD5-RFC clamp unloader after the ligation of the nascent strands (Figure 1) (60,61). This constant loading and unloading, suggests that chromatin bound PCNA levels exist in a “Goldilocks zone” that must be properly maintained. Perturbances to this balance, such as failure to remove PCNA from the chromatin, leads to an increase in recombination and genome instability (45). Furthermore, specific post-translational modifications such as phosphorylation and acetylation can change the levels of chromatin-bound PCNA and result in promiscuous DNA repair activity (62–64). Thus, altering the balance of chromatin-bound PCNA can interfere with various metabolic and regulatory pathways and profoundly affect genome stability.

The thermostability defect of PARD variants lowers PCNA levels in all compartments, but especially chromatin-bound PCNA. The lower chromatin-bound fraction could be due to decreased loading rate or an increased innate dissociation rate. A reduction in the lifetime of PCNA on chromatin may partially explain the differences these mutants have on different DNA metabolic pathways. For instance, DNA synthesis appears to be relatively unaffected in PARD (8), potentially because the replicative polymerases are thought to act almost immediately after PCNA loading (65). However, PCNA-dependent activities that occur later or more slowly, such as DNA repair, are likely more sensitive to PCNA’ s lifetime on DNA. The idea that unstable PCNA variants impair DNA repair is consistent with the finding that substitutions (C81R, E113G, E143K, D150E, and G178S) in yeast PCNA that reduce chromatin-bound PCNA levels cause defects in repair and the DNA damage response (33,44,46,66). Future experiments will test whether the PARD variants have defects in their residence time on DNA.

### Potential therapeutic applications and prospects for PARD

Our biophysical characterization of PCNA-C148S opens new avenues for potential development of PARD therapeutics. Because both PARD variants have a thermostability defect, increasing either their stability and/or lifetime on DNA may improve PCNA function. Small molecules that stabilize PCNA could be considered a therapeutic avenue. However, this strategy may be challenging because small molecules could interfere with secondary binding sites that are involved in partner interactions with PCNA. Indeed, many drugs that bind to sliding clamps interact with the partner binding cleft and block partner binding (2,67,68). An alternative therapeutic approach for PARD therapy is to increase the levels or lifetime of the chromatin-bound PCNA. This strategy could be implemented at the preventative care level by limiting the patients’ exposure to conditions that dramatically impact their PCNA levels. For instance, UV-induced DNA damage stimulates the mono-ubiquitination of PCNA (69), which enhances the activity of error-prone DNA polymerases (70,71), and also triggers PCNA removal by ATAD5-RFC (72,73). Therefore, it may be advantageous from both a cellular and molecular level to limit the amount of direct sunlight these patients experience. If loss of chromatin-bound PCNA is the true driver of PARD, we envision that inhibiting the ATAD5-RFC clamp unloader may increase PCNA levels on DNA and alleviate PARD symptoms. In addition, the temperature sensitivity of PARD suggests that close temperature monitoring of patients and fever-reduction strategies may decrease disease progression.

Beyond therapeutics, our work highlights the need for increased global sequencing of the *PCNA* gene to understand how mutations can impact human health. The identification of a second disease-causing PCNA allele suggests that there are potentially other PARD-causing variants that have yet to be discovered. Identifying these variants could provide a deeper understanding of the cellular pathways compromised in PARD patients. Furthermore, developing mouse models with each of the PARD-associated variant could illuminate the genetic pathways that lead to a pathophysiological phenotype. Ultimately, understanding how disease-causing variant impacts PCNA function will likely provide new mechanistic insight and lead to the development of effective therapies for PARD

## Supporting information

C148S_structure

## Data Availability

All data produced in the present work are contained in the manuscript

## Acknowledgments

The authors would like to thank the families for their willingness to participate in this study. They also thank Dr. M. Orzalli for donating additional healthy donor cells. Drs. N. Rhind, S. Cantor, W. Royer, and C. Peterson are recognized for their input on the finalization of this manuscript. The authors would like to thank J. Pajak and D. Stafforini for their critical analysis of the manuscript’ s written text. This work was funded by the ACS (RSG-16-230-01), FAPESP (grant 2019-19435-3 São Paulo, Brazil), CNPq (grant 30888/2018-8), CAPES (Financial code 01, Brasília, Brazil), FAPESP (grants 2013/03236-5, 2015 26980-7, 2018/10893-6. CNPq (grant 303294/2020-5). The NCI supported J.M. through the F31 NRSA (5F31CA254328).

## AUTHOR CONTRIBUTIONS

A.A.L.J., F.K., and E.K.L. identified the patients. T.K.H. and A.A.L.J are responsible for organizing the clinical data. B.L.F and A.M.L cultured the cells and performed the bioinformatic analysis on the C148S allele, respectively. J.M., and B.A.K designed all the structural, enzymatic, and cellular biochemistry experiments. B.P., and C.G., grew the PCNA-C148S crystals and data/model building was performed by J.M. J.M performed all the enzymatic and cellular biochemistry experiments. Cellular phenotypes experiments were designed and performed by D.J.M., J.B., V.M., and C.F.M.M. Most of the paper was written by J.M and B.A.K, A.A.L.J. wrote the clinical descriptions, and all authors gave final comments and approval. The authors declare no conflict of interest.

