## Supplemental_Figures for "PCNA thermosensitivity underlies an Ataxia Telangiectasia-like disorder"

Magrino et al.

**Supplemental Text**

**Detailed Patient Case reports.**

Detailed case reports removed for medRxiv standards.

A

**Patient data in panels A to meet medRxiv standards**

B

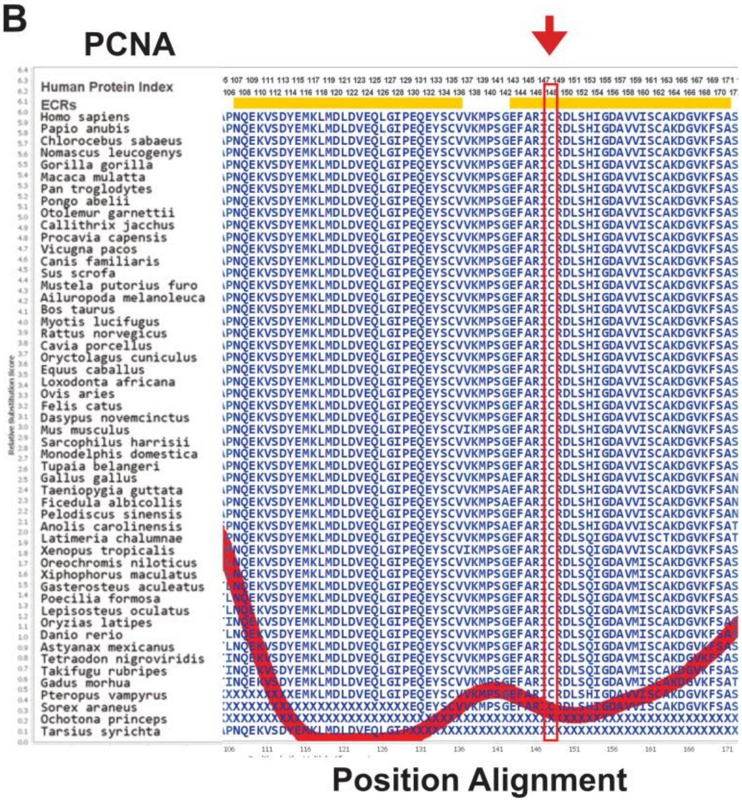


**Supplemental Figure 1**: **Geographic patient distribution and advanced evolution conservation of the C148 allele. (A)** Map of country showing the geographical distribution of the three patients. **(B)** Amino acid alignment of PCNA across 62 vertebrate species depicting the high conservation of C148 (red box, and arrow) using Aminode. The thick red line represents the relative number of substitutions in that region. Areas of low amino acid substitutions are depicted by the thick red line being closer at the bottom of the y-axis (and yellow boxes).

**Supplemental Table 1:** Rare homozygous variants present across the three patients

| Chr | Start | End | Ref | Alt | Gene | Effect on protein | Nucleotide Aminoacid change |
|---|---|---|---|---|---|---|---|
| **Patient 1** | | | | | | | |
| 1 | 152189055 | 152189055 | G | C | HRNR | Nonsynonymous | c.5050C>G:p.Arg1684Gly |
| 1 | 201179497 | 201179497 | G | A | IGFN1 | Nonsynonymous | c.5476A>G:p.Glu1826 Lys |
| 3 | 195510335 | 195510335 | G | A | MUC4 | Nonsynonymous | c.8116T>C:p.Pro2706Ser |
| 4 | 9270413 | 9270413 | T | G | USP17L20 | Nonsynonymous | c.1069G>T:p.Ser357Ala |
| 4 | 9270417 | 9270417 | G | C | USP17L20 | Nonsynonymous | c.1073C>G:p.Ser358Thr |
| 5 | 43280542 | 43280542 | C | T | NIM1K | Nonsynonymous | c.1022T>C:p.Pro341Leu |
| 5 | 96333752 | 96333752 | T | C | LNPEP | Nonsynonymous | c.1556C>T:p.Met519Thr |
| 6 | 30587330 | 30587330 | C | T | MRPS18B | Nonsynonymous | c.139T>C:p.Pro47Ser |
| 6 | 31000073 | 31000074 | AA | - | MUC22 | frameshift deletion | c.4770_4771del:p.Gly1590fs |
| 6 | 36931287 | 36931287 | C | A | PI16 | Nonsynonymous | c.1169A>C:p.Thr390Lys |
| 8 | 10469839 | 10469839 | T | G | RP1L1 | Nonsynonymous | c.1769C>A:p. Gln 590Prp |
| 13 | 50062612 | 50062612 | G | A | SETDB2 | Nonsynonymous | c.1799A>G:p.Arg600Gln |
| 17 | 25973609 | 25973609 | G | A | LGALS9 | Nonsynonymous | c.664A>G:p.Ala222Thr |
| **20** | **5098255** | **5098255** | **C** | **G** | **PCNA** | **Nonsynonymous** | **c.443G>C:p.Cys148Ser** |
| 20 | 37580715 | 37580715 | C | T | FAM83D | Nonsynonymous | c.1310T>C:p.Ser437Phe |
| **Patient 2** | | | | | | | |
| 4 | 5990115 | 5990115 | C | T | C4orf50 | Nonsynonymous | c.1384G>A:p.Ala462Thr |
| 11 | 6633369 | 6633369 | C | T | TAF10 | Nonsynonymous | c.52G>A:p.Ala18Thr |
| 12 | 122958592 | 122958592 | G | A | ZCCHC8 | Nonsynonymous | c.862C>T:p.Arg288Trp |
| 12 | 123109191 | 123109191 | G | A | KNTC1 | Nonsynonymous | c.6562G>A:p.Gly2188Arg |
| 16 | 27221781 | 27221781 | G | T | KDM8 | Nonsynonymous | c.337G>T:p.Ala113Ser |
| **20** | **5098255** | **5098255** | **C** | **G** | **PCNA** | **Nonsynonymous** | **c.443G>C:p.Cys148Ser** |
| **Patient 3** | | | | | | | |
| 9 | 43625382 | 43625382 | G | A | SPATA31A6 | Nonsynonymous | c.C3305T:p.Pro1102Leu |
| 9 | 43627428 | 43627428 | G | A | SPATA31A6 | Nonsynonymous | c.C1259T:p.Pro420Leu |
| 10 | 47000004 | 47000004 | T | C | GPRIN2 | Nonsynonymous | c.T1124C:p.Val375Ala |
| 10 | 51568378 | 51568378 | T | G | NCOA4 | Nonsynonymous | c.T22G:p.phe8Val |
| 10 | 51623190 | 51623190 | T | C | TIMM23 | Nonsynonymous | c.A25G:p.Asn9Asp |
| 16 | 21848694 | 21848694 | T | A | NPIPB4 | Nonsynonymous | c.A1014T:p.Lys338Asn |
| **20** | **5098255** | **5098255** | **C** | **G** | **PCNA** | **Nonsynonymous** | **c.443G>C:p.Cys148Ser** |
| 21 | 10920098 | 10920098 | T | C | TPTE | Nonsynonymous | c.A1156G:p.Lys386Glu |

**Supplemental Table 2.** C148S substitution in-silico pathogenic score

| Program | Score | Cutoff | Prediction |
|---|---|---|---|
| SIFT | 0.03 | < 0.05 | Deleterious |
| PolyPhen2 (HumVar) | 0.888 | > 0.5 | Possibly Deleterious |
| PROVEAN | -7.69 | ≤ -2.5 | Deleterious |
| CADD | 26.7 | < 15 | Deleterious |
| Mutation Assessor | 3.81 | > 0.5 | Deleterious |
| REVEL | 0.91 | ≥ 0.6 | Deleterious |

**Supplemental Table 3:** Data collection and refinement statistics of PCNA-C148S

| **Data Collection** | **PCNA-C148S** |
|---|---|
| Space Group | C 1 2 1 |
| Wavelength | 1.54 |
| Resolution range | 34.97 - 3.10 |
| Unit cell dimensions | |
| a, b, c (Å) | 137.64, 80.87, 70.05 |
| α, β, γ (°) | 90.00, 117.52, 90.00 |
| No. of total reflections | 30,621 (2,294) |
| No of unique reflections | 11,754 (1,059) |
| Multiplicity | 2.6 (2.2) |
| Completeness (%) | 93.0% (83.4%) |
| Mean L/σI | 6.9 (1.5) |
| Wilson B factor | 42.81 |
| $R_{merge}$ | 0.136 (0.608) |
| $R_{meas}$ | 0.169 (0.782) |
| $R_{pim}$ | 0.099 (0.485) |
| $CC_{1/2}$ | 0.983 (0.604) |
| **Refinement** | |
| Resolution | 3.1 Å |
| $R_{work}$ /$R_{free}$ % | 25.52 / 29.21 |
| RMSD | |
| Bonds (Å) | 0.003 |
| Bond angles (°) | 0.764 |
| Ramachandran favored % | 97.92 |
| Ramachandran outliers % | 0.00 |
| Rotamer outliers% | 0.00 |
| Clashscore | 10.54 |
| Average B factor | 40.13 |

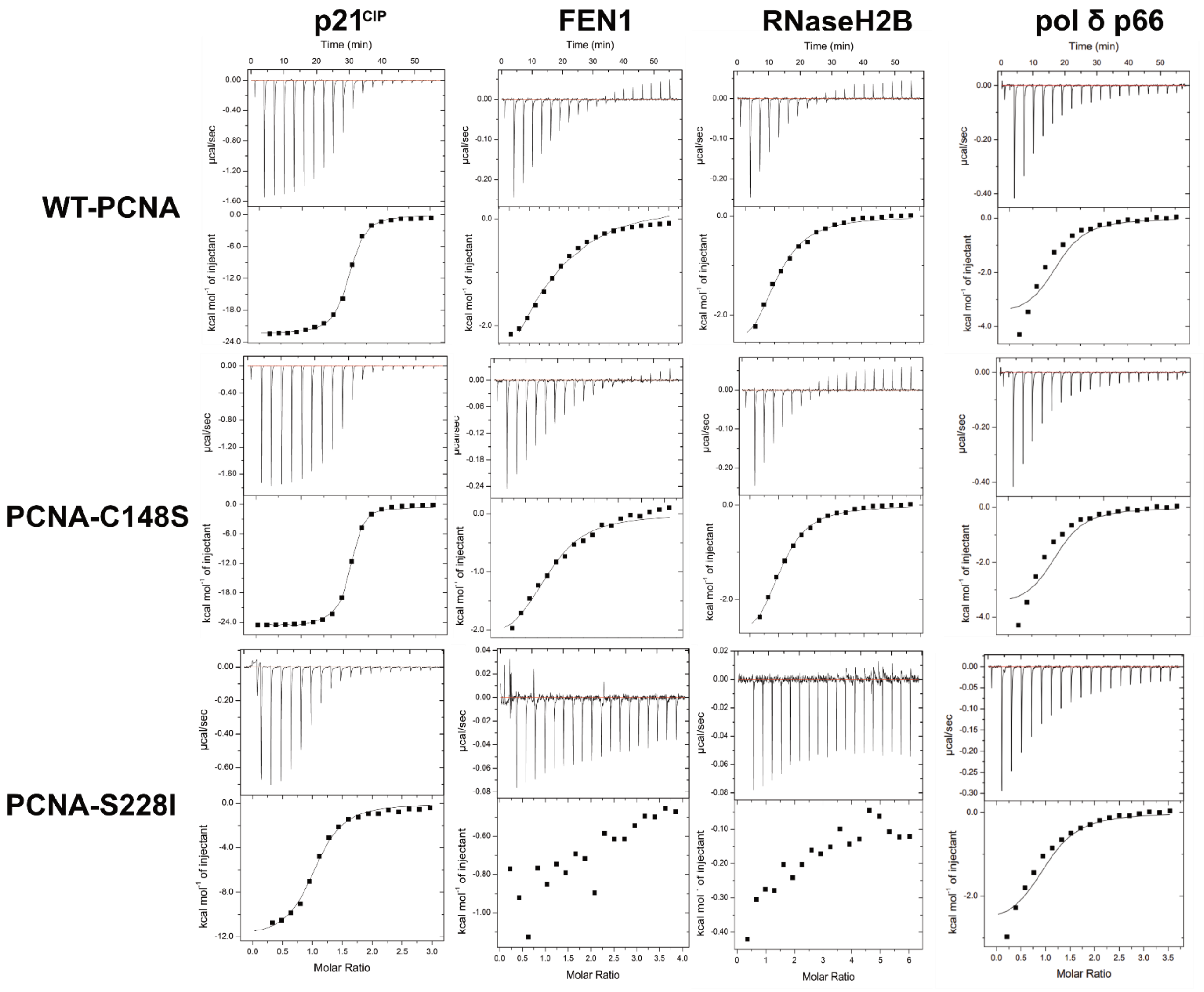


**Supplemental Figure 2: ITC binding curves of various PCNA partner peptides.** Representative examples of the raw ITC binding data for $p21^{CIP}$, FEN1, RNaseeH2B and pol δ for each PCNA variant. See Supplemental Table 4 for the thermodynamic parameters for binding. PCNA-S228I with $p21^{CIP}$, FEN1, RNaseeH2B are from Duffy *et al* 2016.

**Supplemental Table 4:** ITC statistics

| **Protein** | **Ligand** | $K_D$ **(μM)** | $\Delta G$ **(kcal/mol)** |
|---|---|---|---|
| PCNA-WT | $P21^{CIP}$ | 0.21 ± 0.04 | -9.3 ± 0.10 |
| PCNA-C148S | $P21^{CIP}$ | 0.22 ± 0.07 | -9.3 ± 0.20 |
| PCNA-S228I | $P21^{CIP}$ | 0.87 ± 0.07 | -8.7 ± 0.20 |
| PCNA-WT | FEN1 | 11 ± 2.0 | -6.8 ± 0.10 |
| PCNA-C148S | FEN1 | 11 ± 2.0 | -6.7 ± 0.20 |
| PCNA-S228I | FEN1 | Too weak to fit | Too weak to fit |
| PCNA-WT | RNaseH2B | 9.7 ± 0.3 | -6.9 ± 0.03 |
| PCNA-C148S | RNaseH2B | 7.8 ± 0.01 | -7.0 ± 0.01 |
| PCNA-S228I | RNaseH2B | Too weak to fit | Too weak to fit |
| PCNA-WT | P66 | 3.0 ± 0.16 | -7.6 ± 0.05 |
| PCNA-C148S | P66 | 2.8 ± 0.33 | -7.7 ± 0.07 |
| PCNA-S228I | P66 | 4.0 ± 0.14 | -7.5 ± 0.02 |

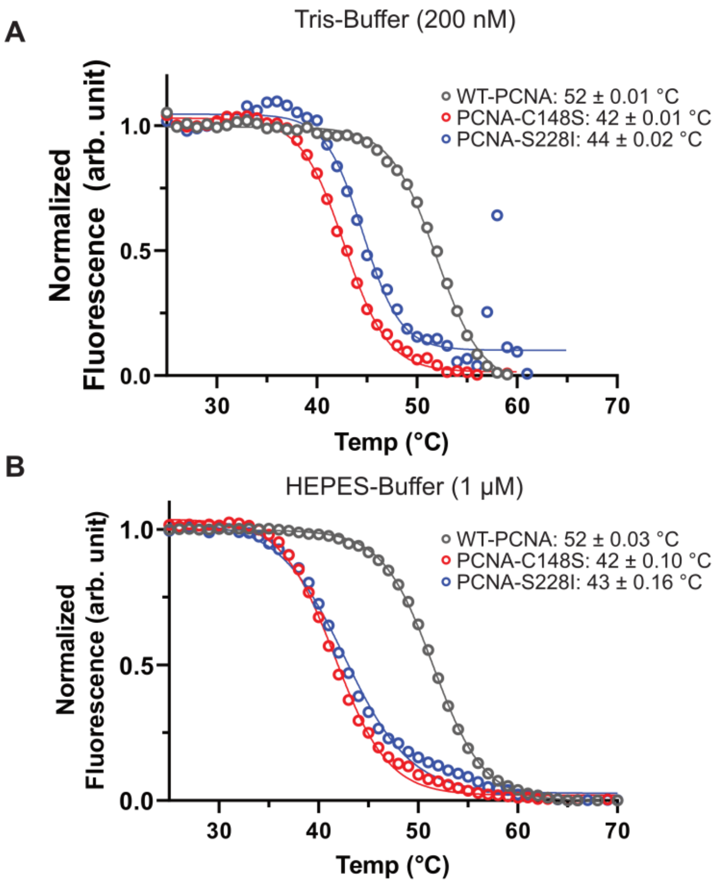


**Supplemental Figure 3: Thermal melts different pH and buffer conditions. (A)** Thermal melt of each variant with 200 nM of PCNA. Each variant displays a two-state curve regardless of protein concentration. **(B)** Thermal melt of each variant in HEPES buffer using 1 µM protein. Both PARD-associated PCNA variants are less stable than WT-PCNA with HEPES buffer.

**Supplemental Table 5.** Unfolding statistics.

| Protein | $T_m$ (C$^o$) | $\Delta G$ (kcal/mol) | $\Delta H$ *(kcal/mol)* | *m* (kcal/mol/[D]) | $C_m$ (M Gdm-HCl) |
|---|---|---|---|---|---|
| PCNA-WT | 52.0 ± 0.03 | 8.20 ± 0.04 | 150 ± 3. | 3.21 ± 0.01 | 2.55 |
| PCNA-C148S | 42.0 ± 0.06 | 7.59 ± 0.60 | 111 ± 3 | 3.73 ± 0.33 | 2.03 |
| PCNA-S228I | 44.0 ± 0.10 | - | 103.6 ± 5 | - | - |

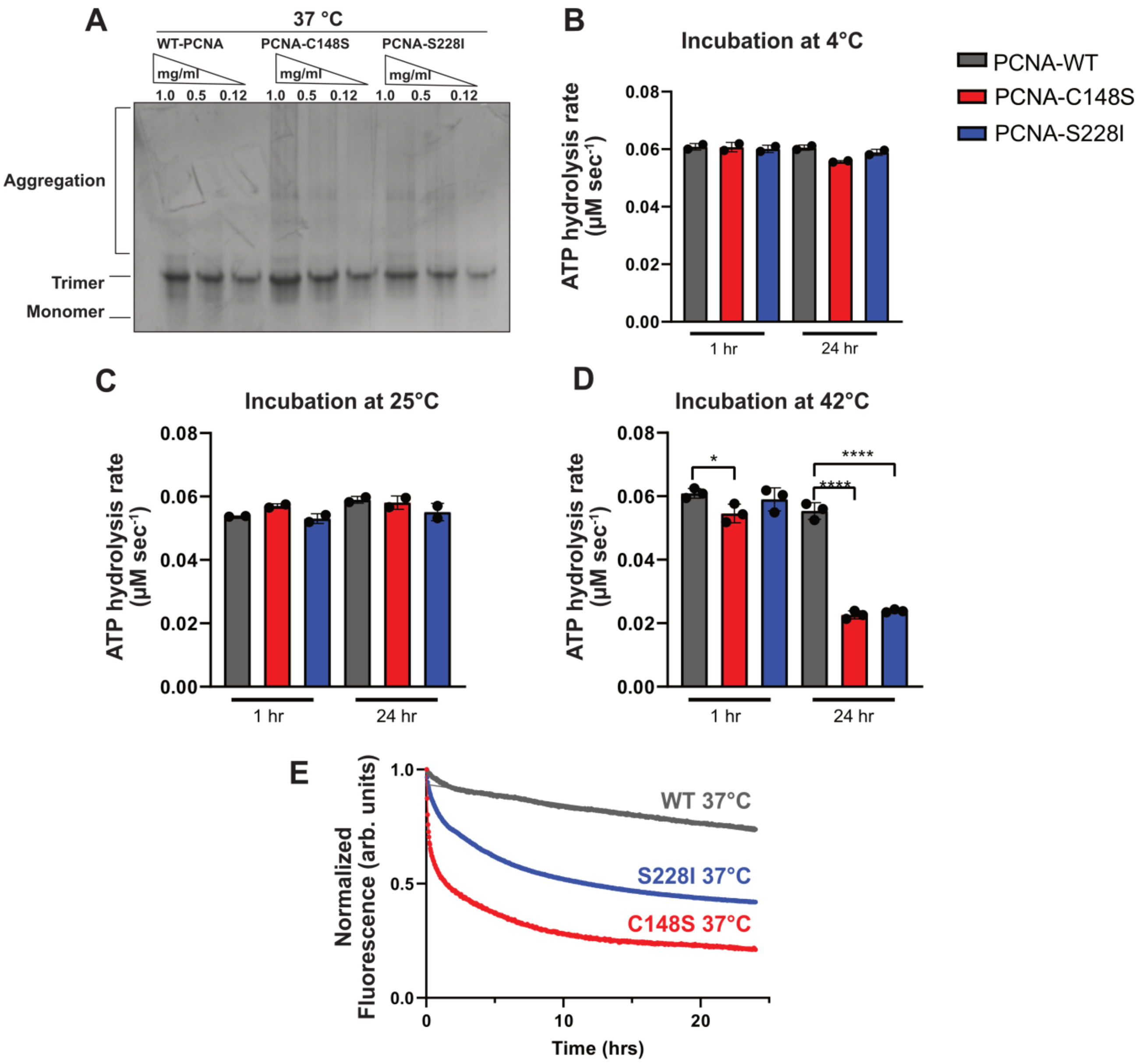


**Supplemental Figure 4: PARD-associated variants are inactivated at elevated temperatures. (A)** Native gel electrophoresis assay to separate trimeric and monomeric PCNA. All variants were incubated at 37 ˚C for 24 hours before electrophoresis. **(B-D)** ATPase assays with protein incubated at various temperatures for 24 hours prior to the ATPase assay conducted at 25 ºC. PARD variants fail to stimulate RFC after a 24-hour pre-incubation at 42 ˚C. **(E)** Tryptophan fluorescence of each variant at 37 ˚C shows that the PARD variants unfold faster than WT-PCNA.

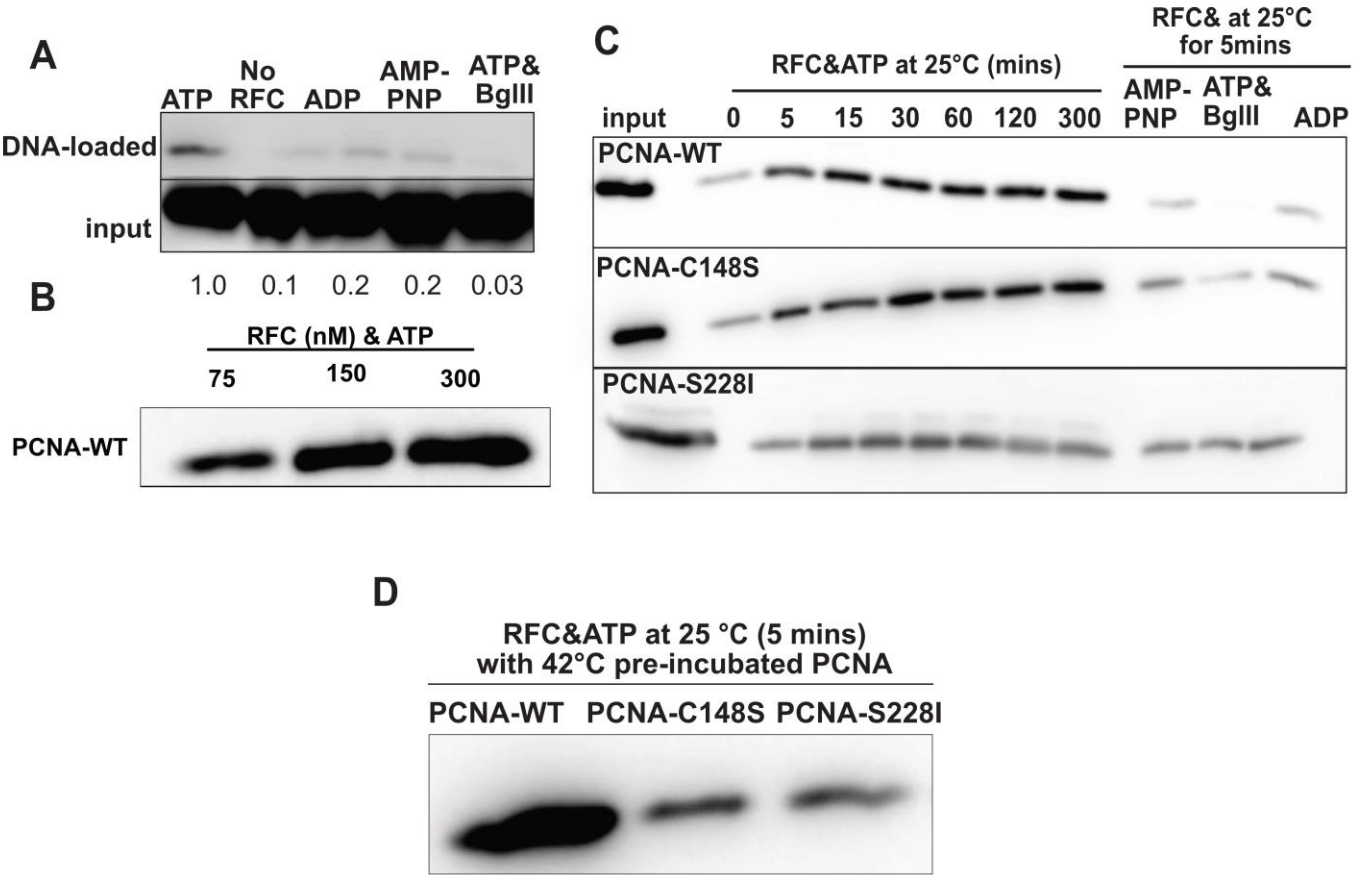


**Supplemental Figure 5: Bead-based PCNA Loading assay. (A)** Representative image of PCNA loading assay with controls, including normalized values relative to an ATP-containing reaction. Loading proceeded for 5 minutes at 25 ºC before addition of stop solution containing EDTA. **(B)** Levels of PCNA loaded onto DNA with increasing concentrations of RFC at 25 ºC for 5 minutes. **(C)** Representative image of PCNA loading kinetics. Time indicates how long each PCNA variant was incubated with RFC at 25 ºC before addition of EDTA stop solution. Controls were all incubated with RFC and conducted for 5 minutes. **(D)** Representative image of PCNA loading conducted at 25 ºC for 5 minutes with protein pre-incubated at 42˚C for 24 hours.

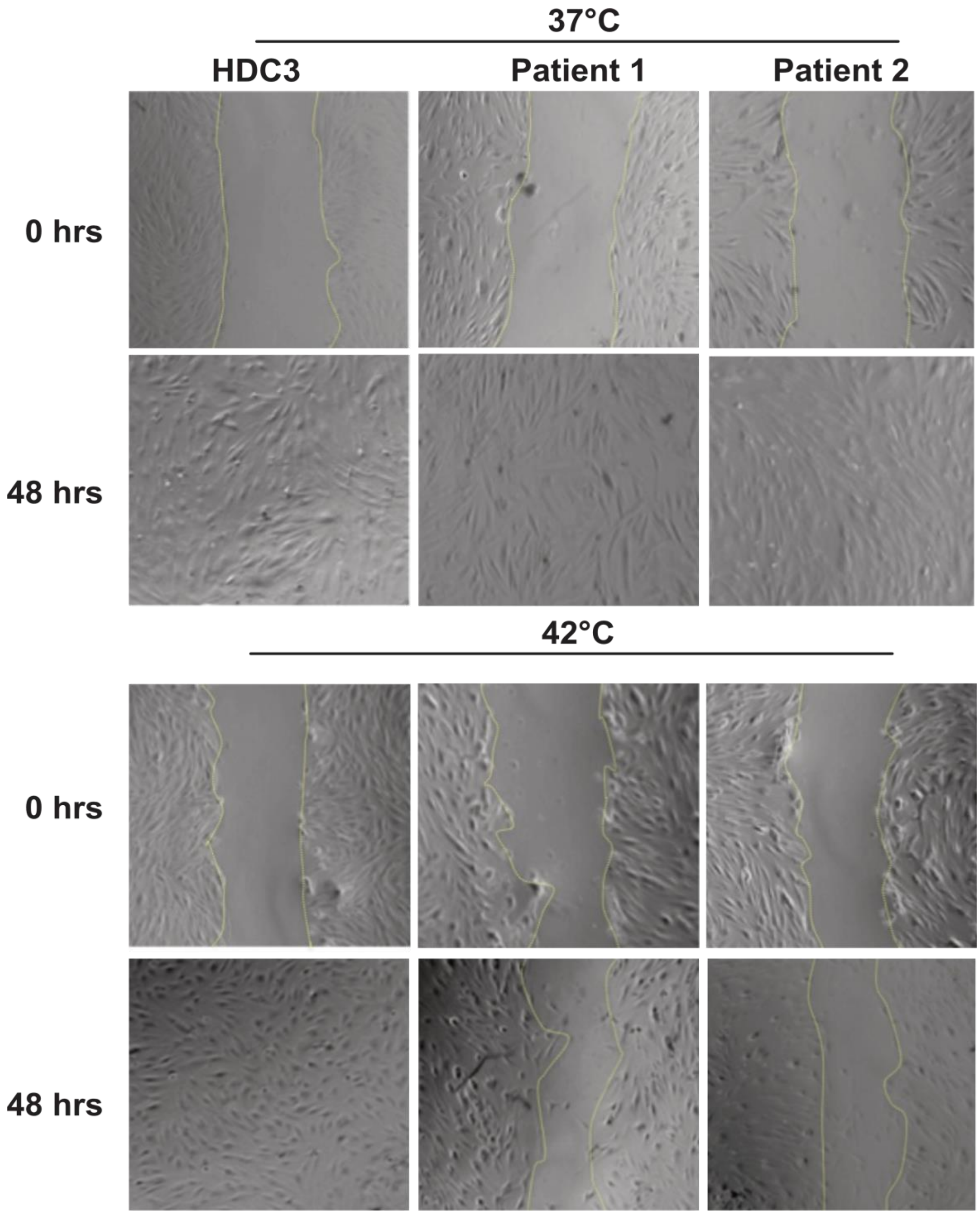


**Supplemental Figure 6: Wound healing assay.** Representative images of patient-derived PCNA-C148S expressing fibroblasts and HDCs incubated at 37 ˚C and 42˚C for 48 hours. At 42 °C, the patient-derived cells exhibit a growth defect.
